# Markers of Biological Brain Aging Mediate Effects of Vascular Risk Factors on Cognitive and Motor Functions: A Multivariate Imaging Analysis of 40,579 Individuals

**DOI:** 10.1101/2024.07.24.24310926

**Authors:** Marvin Petersen, Moritz A. Link, Carola Mayer, Felix L. Nägele, Maximilian Schell, Jens Fiehler, Jürgen Gallinat, Simone Kühn, Raphael Twerenbold, Amir Omidvarnia, Felix Hoffstaedter, Kaustubh R. Patil, Simon B. Eickhoff, Götz Thomalla, Bastian Cheng

## Abstract

The increasing global life expectancy brings forth challenges associated with age-related cognitive and motor declines. To better understand underlying mechanisms, we investigated the connection between markers of biological brain aging based on magnetic resonance imaging (MRI), cognitive and motor performance, as well as modifiable vascular risk factors, using a large-scale neuroimaging analysis in 40,579 individuals of the population-based UK Biobank and Hamburg City Health Study. Employing partial least squares correlation analysis (PLS), we investigated multivariate associative effects between three imaging markers of biological brain aging – relative brain age, white matter hyperintensities of presumed vascular origin, and peak-width of skeletonized mean diffusivity – and multi-domain cognitive test performances and motor test results. The PLS identified a latent dimension linking higher markers of biological brain aging to poorer cognitive and motor performances, accounting for 94.7% of shared variance. Furthermore, a mediation analysis revealed that biological brain aging mediated the relationship of vascular risk factors — including hypertension, glucose, obesity, and smoking — to cognitive and motor function. These results were replicable in both cohorts. By integrating multi-domain data with a comprehensive methodological approach, our study contributes evidence of a direct association between vascular health, biological brain aging, and functional cognitive as well as motor performance, emphasizing the need for early and targeted preventive strategies to maintain cognitive and motor independence in aging populations.

## 1 Introduction

Rising global life expectancy amplifies the challenge of age-related cognitive and motor impairment, threatening functional independence of individuals and burdening societies and healthcare systems worldwide [1]. Although cognitive and motor abilities generally decline with age, there is substantial interindividual variability. While some individuals face cognitive impairment, dementia, and loss of independence, others maintain their cognitive and physical abilities well into advanced age [2]. Unraveling the processes that uphold functional ability in older adults is essential for devising effective prevention and management strategies.

One theory suggests that biological age might diverge from chronological age, potentially offering a more accurate reflection of an individual’s health status in relation to aging [3]. Mechanistic models have thus been proposed, suggesting that interindividual variability in mid and late life functionality arises from variations in biological brain aging, with some individuals exhibiting slower aging processes and others showing accelerated changes [3]. While some contributors to these variations, such as genetic factors, are unmodifiable, others, including lifestyle and environmental influences, are modifiable. Here, cerebrovascular risk factors are considered to contribute to the variation in the rate of biological aging [4].

Indicators of biological brain aging include changes in brain morphology, white matter microstructure, and presence of cerebral small vessel disease (CSVD) [5–8]. Magnetic resonance imaging (MRI) enables to characterize these brain anatomical aspects in vivo. There are multiple neuroimaging markers theorized to capture variation in biological brain aging. These include (1) relative brain age, which measures the the brain age gap – i.e., the discrepancy between chronological age and predicted biological age based on regional brain morphology [9,10], (2) white matter hyperintensities of presumed vascular origin (WMH) indicative of CSVD [11,12], and (3) peak-width of skeletonized mean diffusivity (PSMD) reflecting global microstructural white matter integrity known to decline with increasing age [13].

Despite evidence linking these markers to cognition and motor function, an understanding of the associations remains limited for several reasons [14–16]. Much of the existing research has focused on individual imaging markers, vascular risk factors and cognitive or motor function scores without considering the potentially high covariance of the measures. Moreover, studies often have limited sample sizes, leading to inconsistent results, as emphasized by recent research highlighting the need for larger cohorts to establish reliable links between neuroimaging markers and behavior [17]. Lastly, the role of vascular risk factors in biological brain aging, cognition and motor function is not fully understood, which is particularly relevant given their potential as intervention targets.

To bridge these gaps, we present a large-scale multi-modal neuroimaging analysis in two large-scale epidemiological studies, the UK Biobank (UKB) and the Hamburg City Health Study (HCHS), combining key brain MRI markers of biological brain aging, vascular risk information, and comprehensive cognitive and motor phenotyping with advanced statistics that can aptly capture multivariate and potentially covarying associative effects. In this analysis, we applied data-driven statistics in form of a partial least squares correlation analysis (PLS) to model the multivariate associative effects of biological brain aging, cognition, and motor function. Expanding on this, we investigated the role of biological brain aging in mediating the relationship between vascular risk, cognition and motor function in a mediation analysis. In sum, our analysis aimed to contribute to the understanding of the neurobiology underlying the decline in everyday cognitive and motor functioning to help identify potential diagnostic and treatment targets.

## 2. Material and Methods

An overview of our methodology is provided in *figure 1*. In brief, we utilized imaging and clinical data from the UKB and HCHS to investigate the complex relationships among biological brain aging, modifiable vascular risk, and cognitive and motor function. Following preprocessing, we derived relative brain age, WMH load, and PSMD from the imaging data. We then employed PLS to model the multivariate associations between these imaging markers and clinical scores of cognitive and motor function. Using the resulting subject-level PLS scores, which capture the associative effects between biological brain aging and clinical performance, we conducted a mediation analysis to test whether biological brain aging mediates the relationship between vascular risk measures and cognitive and motor test performance. All these analyses were performed separately in both the UKB (discovery cohort) and HCHS (replication cohort).

**Figure 1.**
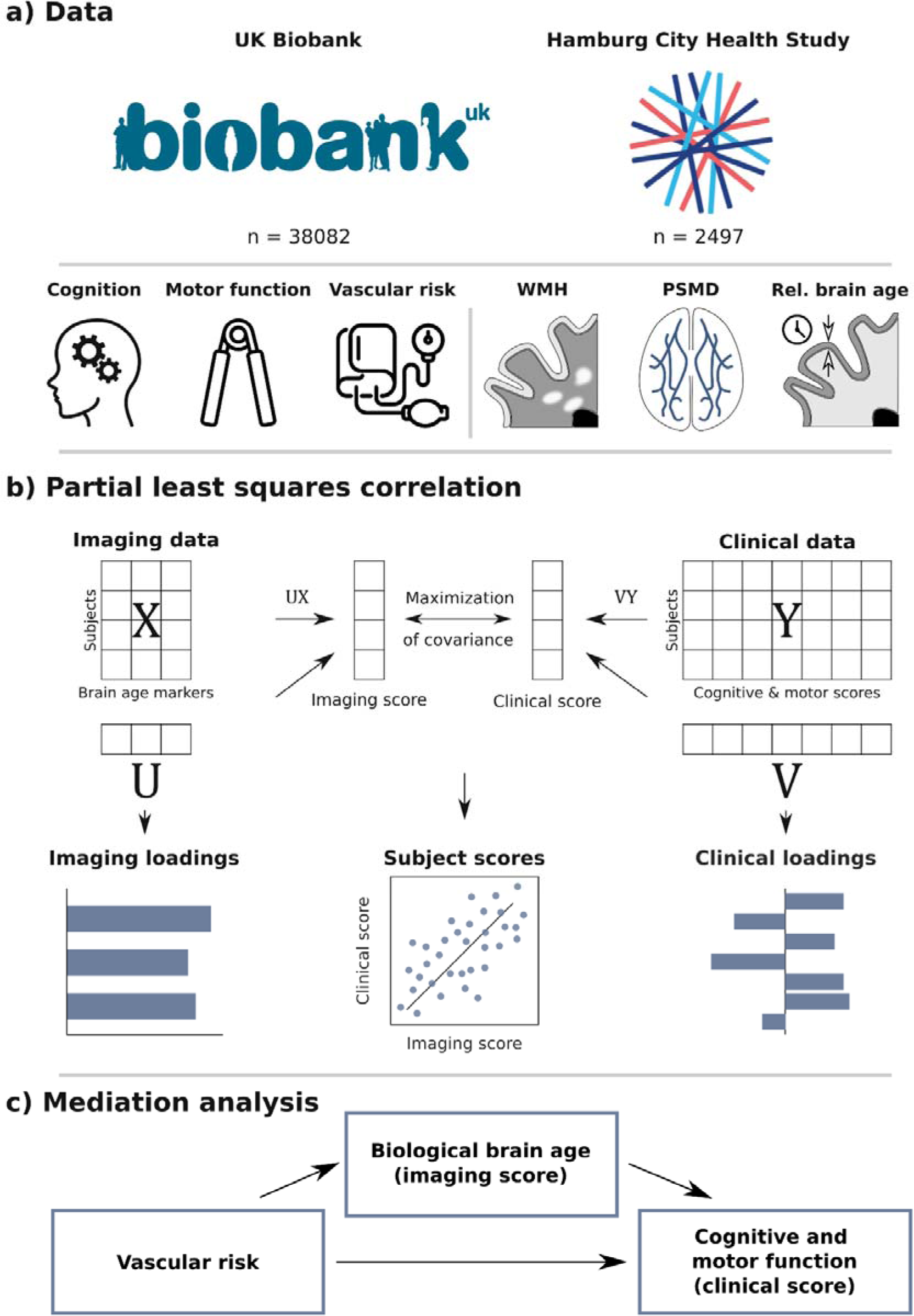
Methodology. a) Population-based data from the UK Biobank and Hamburg City Health Study were used including cognitive test scores, motor performance scores, vascular risk measures and multimodal brain MRI. Three different measures of biological brain aging were derived from anatomical and diffusion-weighted MRI: WMH, PSMD and relative brain age. b) Imaging measures of biological brain age were related to cognitive and motor performance scores via partial least squares correlation analysis (PLS). PLS computes subject-specific scores (here imaging and clinical score), combining input data (X & Y) and respective loadings (U & V) through a linear combination. The loadings reveal the associative impact of the two input data domains, comparable to β-coefficients in linear regression. Together, subject-specific scores and loadings represent a latent variable. c) The interplay between biological brain aging, vascular health, as well as cognitive and motor performance was investigated in a post-hoc mediation analysis. We tested whether the relationship between different vascular risk measures and the clinical score – representing cognitive and motor performance, was statistically mediated by the imaging score – representing biological brain aging. Abbreviations: PSMD – peak width of skeletonized of mean diffusivity, WMH – white matter hyperintensities of presumed vascular origin.

### 2.1 Study population

The presented study is based on behavioral and neuroimaging data from two large-scale population-based cohort studies, the UKB (n=43,098, age range 40-69 years) and HCHS (n=2,652, age range 45-74 years).

### 2.2 Ethics statement

The UKB’s ethical approval was granted by the North West Multi-Centre Research Ethics Committee (MREC). Details on the Ethics and Governance framework are provided online (https://www.ukbiobank.ac.uk/media/0xsbmfmw/egf.pdf) [18]. The HCHS was approved by the local ethics committee of the Landesärztekammer Hamburg (State of Hamburg Chamber of Medical Practitioners, PV5131). Written informed consent was obtained from all participants. Data acquisition procedures followed the Good Clinical Practice (GCP) and Good Epidemiological Practice (GEP) guidelines according to the Declaration of Helsinki [19].

### 2.3 Clinical Data

Measurements of cognitive and motor function were investigated in this work. For more detailed descriptions of the administered cognitive and motor tests used in this study also refer to the *supplementary materials S1*.

In brief, cognitive functioning was assessed in the UKB administering tests for executive functioning (Tower Rearranging Test, Trail Making Test B), processing speed (Reaction Time Test, Symbol Digit Substitution Test, Trail Making Test A), memory (Numeric Memory Test, Paired Associate Learning Test), and reasoning (Fluid Intelligence Test, Matrix Pattern Completion Test). In the UKB, cognitive tests were administered as computerized test versions. Motor capacity was characterized via hand grip strength measurements of right and left hand and evaluation of accelerometry as an indicator of physical activity [20].

In the HCHS, cognitive testing covered executive functioning (Trail Making Test B), processing speed (Trail Making Test A), memory (Word List Recall Test), reasoning (Multiple Choice Vocabulary Intelligence Test B) and language (Animal Naming Test). Motor function was examined via hand grip strength measurements of the right and left hand and the Timed Up And Go Test. In the HCHS, tests were administered by trained professionals.

To facilitate interpretability and enhance comparability between examined cohorts cognitive test results were harmonized. The harmonization procedure was based on previous protocols [21]. First, cognitive test scores were assigned to cognitive domains. Within the cognitive domains, the corresponding tests were z-scored and averaged to obtain domain scores. Domain scores were computed for the domains executive functioning, processing speed, memory and reasoning for the UKB and executive functioning, processing speed, memory, reasoning, and language for the HCHS. If multiple tests were available for a specific cognitive domain, their z-scores were averaged. Trail Making Test and reaction time results were inverted and log transformed beforehand according to previous work [20]. Furthermore, grip strength of the left and right hand was averaged. Mean hand grip strength was normalized based on division by the square of individual height following previous procedures [22].

### 2.4 Image processing

MRI acquisition protocols, preprocessing and quality assessment are documented in detail in *supplementary materials S2*. Three measures of biological brain aging were derived from MR images: relative brain age, WMH load and PSMD. For the UKB, precomputed data were used, if possible. This included morphometric data computed via FreeSurfer, WMH segmentations as well as preprocessed diffusion-weighted imaging data. HCHS data was fully processed by our team.

#### 2.4.1 Brain morphometry and relative brain age

Estimation of cortical and subcortical volumetric indices was performed leveraging Freesurfer. Technical details of the comprised procedures were described before [23–25]. In brief, the processing included motion correction, intensity normalization, removal of non-brain tissue, segmentation of subcortical structures and cortical surface reconstruction. Cortical thickness was measured on the vertex level as the distance between the pial surface and grey matter-white matter-boundary [26]. Cortical thickness and subcortical volumes were aggregated within regions of interest defined by the Desikan-Killiany cortical atlas and the aseg subcortical atlas [25,27].

Subsequently, region of interest-level morphological measures were used for relative brain age estimation. Notably, the estimation of relative brain age can be based on various imaging modalities. We chose to implement it using regional brain morphology, as this method is both common and computationally feasible for the scale of our study [10]. Hence, the implemented measure captures the morphology of an individual brain in comparison to the population average: a positive relative brain age score represents advanced biological brain age, i.e., an “older appearing brain” compared to age-matched peers, whereas a negative relative brain age score indicates a relatively younger biological brain age [28]. Importantly, relative brain age represents a variation of the commonly used brain age gap, i.e., the difference between chronological age and the predicted biological age from neuroimaging data (*predicted age*), also referred to as brainAGE or brain predicted-age difference score (brainPAD) [10]. Due to regression dilution bias, this original brain age gap measure can be negatively correlated with chronological age [28,29]. Relative brain age addresses this ensuring orthogonality with chronological age [9,29]. Computations were performed leveraging julearn (v. 0.2.4) and scikit-learn (v. 0.24.1) [30]. First, *predicted age* was obtained by fitting an ordinary least squares regression model with cortical thickness and subcortical volumetric indices as features and chronological age as the target [23,25]. Fitting procedures were performed in a 5-fold cross-validation (see *supplementary table S3* for prediction scores). In a second step, *expected age* was determined by fitting a separate linear model, but this time using chronological age as the input feature and *predicted age* as the target. Finally, relative brain age was calculated as the difference between predicted and expected brain age.

Relative Brain Age = Predicted Age - Expected Age

In accordance to previous procedures, predictions were performed separately for male and female participants [31]. For regression plots displaying orthogonality between relative brain age and chronological age see *supplementary figure S4*.

#### 2.4.2 White matter hyperintensity segmentation

White matter hyperintensities of presumed vascular origin (WMH) indicate regions with severe age-related small vessel pathology. WMH segmentation was performed by applying FSL’s Brain Intensity AbNormality Classification Algorithm (BIANCA) – a fully automated, supervised machine learning approach for WMH detection based on k-nearest neighbor classification – to FLAIR and T1w images [32]. WMH load was calculated as the ratio of WMH volume to intracranial volume computed via FreeSurfer and logarithmized to ensure a normal distribution following previous procedures [33].

#### 2.4.3 Peak width of skeletonized mean diffusivity

The PSMD is a global marker of microstructural white matter integrity [34]. PSMD was calculated via diffusion tensor imaging based on preprocessed diffusion-weighted images. First, diffusion tensors were modelled via a least-squares fit. From the resulting tensors, mean diffusivity maps were derived. Skeletonized maps of mean diffusivity were obtained following the tract-based spatial statistics (TBBS) procedure [35]. PSMD was calculated as the difference between the 95^th^ and 5^th^ percentiles of the MD voxel values within the skeleton [33].

### 2.6 Statistics

Visualization and statistical analysis were performed using Python (v. 3.9.7) leveraging julearn (v. 0.2.4), matplotlib (v. 3.3.4), numpy (v. 1.2.1), pyls (v. 0.0.1), pandas (v. 1.2.4.), pingouin (v. 0.5.0), scikit-learn (v. 0.24.1), seaborn (v. 0.11.1), statsmodels (v. 0.13.1), confounds (v. 0.1.3), proplot (v. 0.9.5). Results were considered significant at a p-value of < 0.05. To address multiple testing, reported p-values were false discovery rate-corrected. Descriptive statistics of the UKB and HCHS involved calculation of mean and standard deviations.

#### 2.6.1 Partial least square correlation analysis

PLS was leveraged to investigate the multivariate associative effects between imaging and behavioral markers. Prior to PLS, imaging and clinical variables were residualized against chronological age, sex and education for deconfounding. Associations between imaging and clinical variables to age before residualization can be found in *supplementary figures S5 and S6*. PLS was performed via pyls (https://github.com/rmarkello/pyls). For a detailed methodological description of PLS please refer to *supplementary text S7* [18]. In brief, PLS identifies associative effects between two sets of variables by identifying latent variables maximizing their covariance. In case of the presented study, the relationship of imaging markers of brain aging (relative brain age, WMH load, PSMD) and clinical data (cognitive function, motor test results and chronological age, sex, education) was modeled. A latent variable consists of a singular value as well as loadings for both input domains respectively quantifying the contribution of imaging and clinical variables to the overall covariance profile represented by the latent variable. In a simplified perspective, PLS can be considered as a dual regression resulting in interpretable coefficients for both multivariable data domains – i.e., a many-to-many mapping instead of a many-to-one mapping as provided by β coefficients in multiple regression. Significance testing of latent variables was performed by permuting subject labels of the imaging data domain and comparison of empirical singular values to the permuted distribution (n_permutation_ = 5000). Robustness of individual singular vector loadings was assessed via bootstrap resampling (n_bootstrap_ = 5000). Bootstrapping involves random resampling with replacement, yielding a distribution of loadings for each variable. This enables the computation of 95% confidence intervals for the clinical variables and a bootstrap ratio 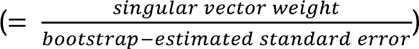 for the brain imaging markers. The bootstrap ratio measures the robustness of a brain imaging markers contribution to the observed covariance pattern. A region with a robust contribution demonstrates a high loading alongside a small standard error, i.e., stability across bootstraps. For clinical variables, a confidence interval excluding 0 (0 ∉ [Confidence interval]) indicated significance. For imaging markers, a bootstrap ratio exceeding 1.96 or less than −1.96 for imaging markers was considered as significant. Furthermore, subject-level imaging and clinical PLS scores were calculated quantifying the extent an individual expressed the identified imaging or clinical covariance profile. For instance, a higher imaging score indicates a higher adherence of a participant to the respective differences in imaging markers of brain aging [5].

To enhance comparability with previous studies, we supplemented a linear regression analysis of the imaging and clinical variables associated in the PLS. Therefore, relative brain age, WMH load and PSMD were individually related to the cognitive and motor variables in linear regression analyses. All models were adjusted for chronological age, sex and education. Effect sizes were reported as standardized β estimates.

Unlike relative brain age computations, WMH load and PSMD calculations are absolute measures and do not quantify deviations from the population mean. To investigate the stability our results, we derived measures that quantify these deviations for WMH load and PSMD, aligning with the relative brain age concept. Essentially, we computed relative brain age measures but based on WMH load and PSMD instead of regional brain morphology to reflect deviations in CSVD burden and white matter microstructure. We then included these gap measures – termed WMH brain age and microstructural brain age – in place of the original absolute measures, alongside the initial relative brain age, in the PLS analysis.

#### 2.6.2 Mediation analysis

To disentangle the complex interplay between vascular risk factors, cognitive function and motor performance, we performed a post-hoc mediation analysis enabling the examination of biological brain aging as a potentially relevant intermediary in this link [36]. For this analysis, we used the subject-level imaging score and clinical score resulting from the PLS. These scores can be interpreted as summary measures like factors or principal components from other dimensionality reduction techniques: The subject-level imaging score represents a data-driven summary measure of biological brain aging markers while the subject-level clinical score summarizes cognitive and motor performance. We tested the mediating effect of the subject-level imaging score on the association of vascular risk factors and the subject-level clinical score. The considered vascular risk factors included arterial hypertension (systolic and diastolic blood pressure), dyslipidemia (blood levels of cholesterol, HDL, LDL, triglycerides) and glycemia (blood glucose), obesity (waist-hip-ratio) and smoking (pack years). A mediation analysis decomposes the total effect of the vascular risk factors on the subject-level clinical score into two components: (1) the direct, i.e., non-mediated, effect of vascular risk on the clinical score, and (2) the indirect effect, i.e., the proportion of the effect that can be attributed to the subject-level imaging score. An indirect effect was considered to mediate the relationship between vascular risk and clinical performance when a vascular risk factor was significantly associated with the mediator, the mediator was significantly associated with the subject-level clinical score and the link between a vascular risk factor and the subject-level clinical score was reduced (partial mediation) or became non-significant (full mediation) when controlling for the mediator. The presence of a significant mediating effect was determined using bootstrapping (n_bootstrap_=5000). Models were adjusted for chronological age, sex and education. Input variables to the mediation analysis were z-scored beforehand, so standardized effect measures are reported.

## 3 Results

The main manuscript reports results for the UKB. For details on HCHS results refer to the *supplementary materials*.

### 3.1 Descriptive statistics of UKB and HCHS

Application of exclusion criteria and quality assessment resulted in the exclusion of n=5,016 subjects in the UKB sample and n=155 subjects in the HCHS sample. The final analysis sample consisted of n=38,082 UKB subjects and n=2,497 HCHS subjects. For a more detailed overview of the sample selection procedure also refer to *supplementary figure S8*. Descriptive statistics are displayed in ***table 1***.

**Table 1:** Descriptive statistics. Abbreviations: ISCED = International Standard Classification of Education, kg = kilogram, l = liter, mm = millimeters, mmHg = millimeter mercury, sec = seconds, ^a^Presented as mean ± standard deviation.

| **Demographics and vascular risk factors** | **UKB (n = 38,082)<sup>a</sup>** | **HCHS (n = 2,497)<sup>a</sup>** |
| --- | --- | --- |
| Age, years | 63.7 ± 7.6 | 63.79 ± 8.3 |
| Sex, % female | 50.2 | 44.13 |
| Education, ISCED | 4.5 ± 1.3 | 2.43 ± 0.6 |
| Pack years | 18.8 ± 15.7 | 8.09 ± 17.4 |
| Waist-hip ratio | 0.9 ± 0.1 | 0.94 ± 0.09 |
| RR<sub>systolic</sub>, mmHg | 138.6 ± 18.6 | 141.41 ± 19.5 |
| RR<sub>diastolic</sub>, mmHg | 78.7 ± 10 | 82.64 ± 10.1 |
| Cholesterol, mmol/l | 5.7 ± 1.1 | 5.43 ± 1.1 |
| LDL, mmol/l | 3.6 ± 0.8 | 3.15 ± 1 |
| HDL, mmol/l | 1.5 ± 0.4 | 1.66 ± 0.5 |
| Triglycerides, mmol/l | 1.6 ± 1 | 1.36 ± 0.8 |
| Glucose, mmol/l | 5.0 ± 1 | 5.38 ± 1.1 |
| **Imaging markers** |  |  |
| Relative brain age | -0.01 ± 3.8 | -0.001 ± 4.6 |
| PSMD, 10<sup>-4</sup> mm<sup>2</sup>/s | 2.29 ± 0.4 | 2.26 ± 0.3 |
| WMH load, % | 0.29 ± 0.4 | 0.17 ± 0.2 |
| WMH volume, ml | 4.63 ± 5.5 | 2.56 ± 3.2 |
| **Cognitive tests** |  |  |
| Numeric Memory Test | 6.79 ± 1.3 |  |
| Trail Marking Test A, s | 22.45 ± 8.1 | 39.92 ± 14.3 |
| Trail Marking Test B, s | 57.13 ± 25.7 | 89.26 ± 38.3 |
| Matrix Pattern Completion Test | 8.03 ± 2.1 |  |
| Fluid intelligence | 6.66 ± 2.1 |  |
| Reaction time, s | 0.59 ± 0.1 |  |
| Paired Associate Learning Test | 6.98 ± 2.6 |  |
| Tower Rearranging Test | 9.97 ± 3.2 |  |
| Symbol Digit Substitution Test | 19.06 ± 5.2 |  |
| Word List Recall Test |  | 7.77 ± 1.8 |
| Multiple Choice Vocabulary Intelligence Test B |  | 31.26 ± 3.6 |
| Animal Naming Test |  | 24.85 ± 6.9 |
| **Motor tests** |  |  |
| Mean hand grip strength, kg | 30.3 ± 10.3 | 34.26 ± 10.6 |
| Average acceleration, milli-gravity | 28.5 ± 7.8 |  |
| Timed Up And Go Test (s) |  | 7.02 ± 1.7 |

### 3.2 Imaging markers of biological brain aging are associated with cognitive and motor function

PLS was performed to map the multivariate associative effect pattern of imaging markers of biological brain aging, motor and cognitive performance measures within a single model. PLS revealed three significant latent variables (*figure 2a*) each representing a many-to-many mapping relating imaging and clinical markers. The first latent variable accounted for 94.7 % of shared variance and was thus further examined.

**Figure 2.**
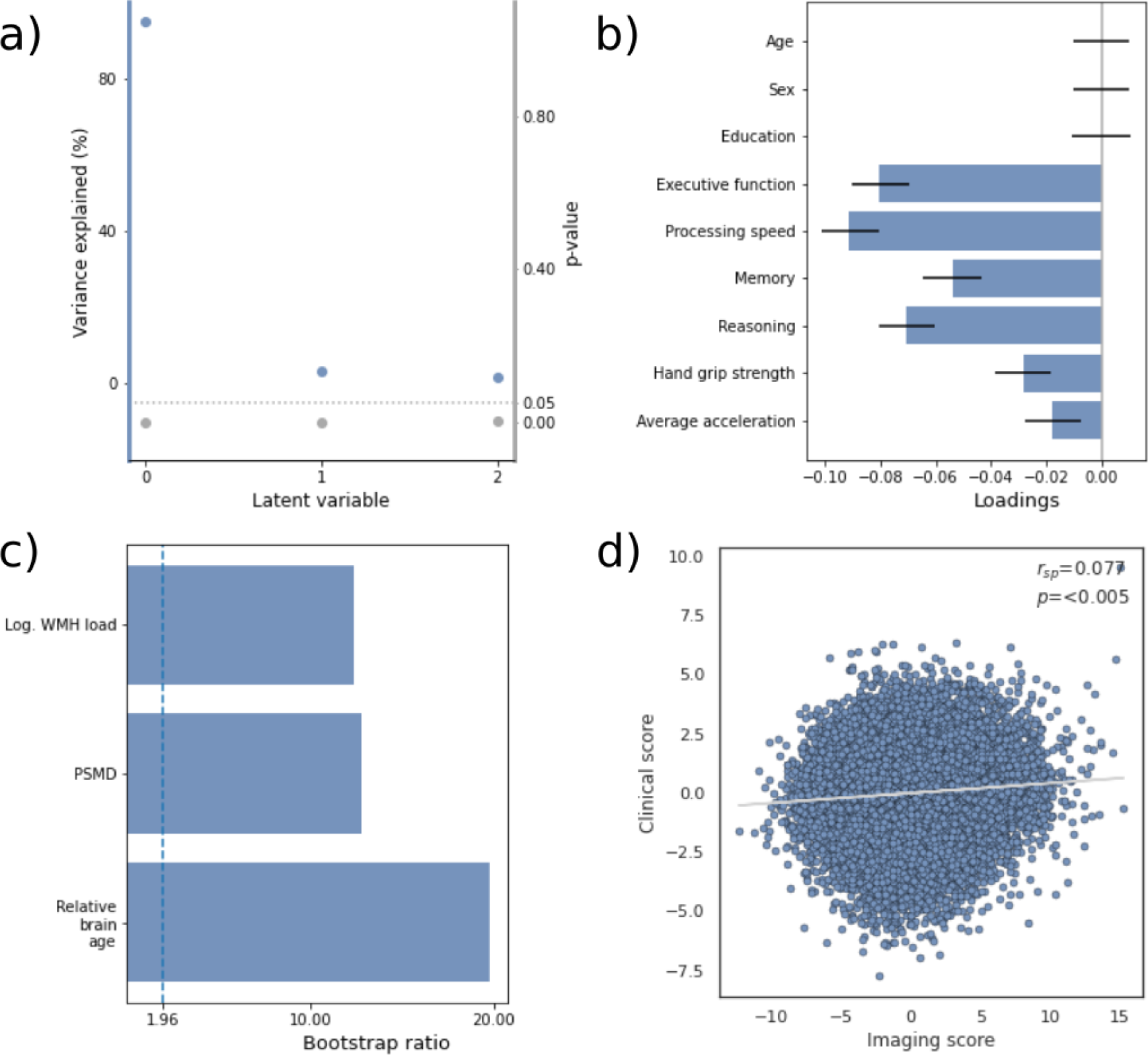
Partial least squares correlation results. a) Overview of detected latent variables with the first latent variables explaining 94.7% of observed shared variance. b) Loadings of the clinical variable set consisting of cognitive and motor test results. Error bars indicate the 95% confidence interval obtained by bootstrap resampling. c) Bootstrap ratios of the imaging variable set. The vertical dashed line represents the significance threshold (bootstrap ratio > 1.96). d) Relationship of subject-level imaging and clinical scores. Abbreviations: r_sp_ – Spearman correlation, Log. WMH load – logarithmized white matter hyperintensity load, PSMD – peak width of skeletonized mean diffusivity.

Specifically, the first latent variable corresponded with a clinical covariance pattern of significantly worse cognitive and motor performance across all considered clinical variables (0 ∉ [95% confidence interval]; *figure 2b*), i.e., executive function, processing speed, memory, reasoning, hand grip strength and average acceleration (for details see *supplementary table S9*). Notably, cognitive domain scores of executive function and processing speed showed the strongest contribution to the covariance profile as indicated by the highest loading to the latent variable. Chronological age, sex and education did not significantly contribute to the covariance pattern (0 ∈ [95% confidence interval]) indicating sufficient effects of deconfounding. Regarding the imaging markers of biological brain aging, relative brain age ([boostrap ratio], 19.8), PSMD (12.8) and WMH load (12.4) exhibited a significant positive (>1.96) contribution to the covariance pattern (*figure 2c*). Therefore, a higher relative brain age, WMH load and PSMD corresponded with worse cognitive and motor performance. Of note, relative brain age contributed most strongly among investigated imaging markers as indicated by the highest bootstrap ratio. Subject-specific imaging and clinical scores were calculated that illustrate the subject-specific expression of the respective covariance profiles. For instance, a participant with a higher positive imaging score exhibits a higher agreement with the imaging covariance profile – i.e., higher relative brain age, WMH load and PSMD – and vice versa. Per definition, the scores are correlated (T_sp_ = 0.077, p <0.05, *figure 2d*) indicating that individuals expressing the clinical covariance pattern (worse cognitive and motor performance) also express the imaging pattern (higher relative brain age, WMH load, PSMD). This relationship was robust across a 10-fold cross-validation (avg. T_sp_ = 0.096 *supplementary table S10*). The PLS results remained stable when incorporating brain age gap measures based on WMH load and PSMD alongside relative brain age (see *supplementary figure S11*). A PLS including all individual cognitive scores instead of domain scores is shown in *supplementary figure S12*.

In addition, we performed multiple linear regression analyses between individual imaging markers of biological brain aging as well as cognitive and motor performances confirming the associations suggested by the PLS. For multiple linear regression analysis results of the associations between individual clinical and imaging markers please refer to *supplementary materials S13* and *S14*. Cross correlation matrices of imaging and clinical markers are shown in *supplementary materials S15* and *S16*. The abovementioned results were reproducible in the HCHS sample. For details on the HCHS results refer to *supplementary materials S17 – S25*.

### 3.3 Mediation analysis

To investigate whether subject-level imaging scores mediate the relationship between vascular risk factors and subject-level clinical scores, we performed a post-hoc mediation analysis (*figure 3*). We found the subject-level imaging scores to partially mediate the link between vascular risk factors and clinical scores. This held true for systolic blood pressure (ab = 0.004, p_FDR_ < 0.001; c’ = 0.025, p_FDR_ < 0.001; c = 0.029, p_FDR_ < 0.001), diastolic blood pressure (ab = 0.005, p_FDR_ < 0.001; c’ = 0.036, p_FDR_ < 0.001; c = 0.041, p_FDR_ < 0.001), triglycerides (ab = 0.003, p_FDR_ < 0.001; c’ = 0.065, p_FDR_ < 0.001; c = 0.068, p_FDR_ < 0.001), glucose (ab = 0.006, p_FDR_ < 0.001; c’ = 0.020, p_FDR_ < 0.001; c = 0.026, p_FDR_ < 0.001), waist-hip ratio (ab = 0.009, p_FDR_ < 0.001; c’ = 0.126, p_FDR_ < 0.001; c = 0.135, p_FDR_ < 0.001) and pack years (ab = 0.008, p_FDR_ < 0.001; c’ = 0.064, p_FDR_ < 0.001; c = 0.072, p_FDR_ < 0.001). The links between the subject-level cholesterol, HDL cholesterol, LDL cholesterol and the clinical score were not significantly mediated. These results were reproducible in the HCHS except for systolic and diastolic blood pressure showing no significant link to the subject-level clinical score (*supplementary figure S26*).

**Figure 3.**
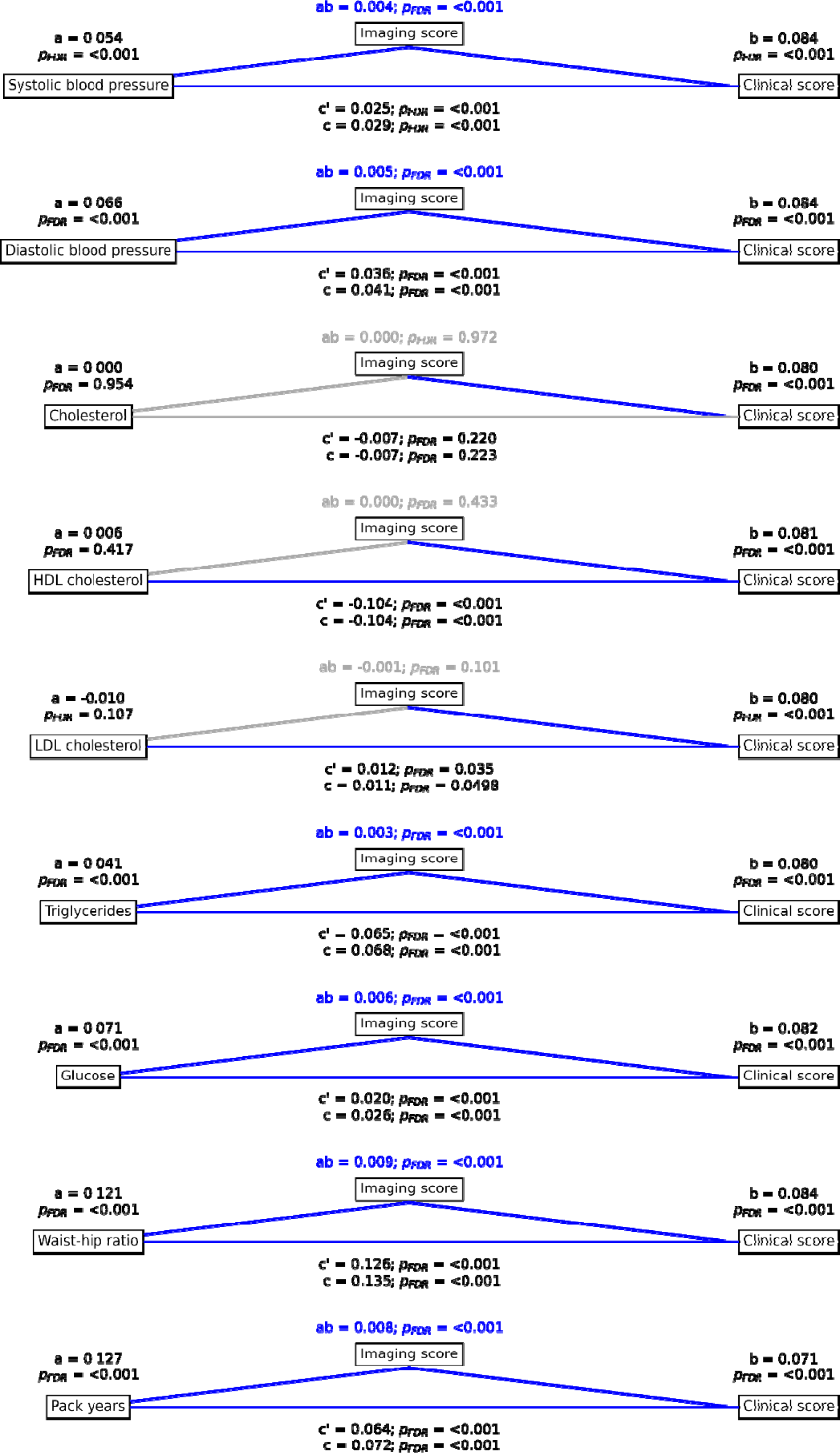
Mediation analysis results. Mediation effects of subject-level imaging score on the relationship between vascular risk factors and subject-level clinical scores summarizing cognitive and motor performance. Path plots display standardized effects and p-values: (a) vascular risk factor to subject-level imaging score, (b) subject-level imaging sore to clinical score, (ab) indirect effect (c’) direct effect and (c) total effect. Significant paths are highlighted in blue; non-significant in light gray. If the indirect path was significant the text for ab is highlighted in blue. Abbreviations: p_FDR_ – false discovery rate-corrected p-value.

## 4 Discussion

Understanding individual differences in aging trajectories is vital for informing strategies to maintain physical and cognitive health in mid and later life. In this work, we linked structural neuroimaging markers of biological brain aging with cognitive and motor test performances in two population-based samples with a total of 40,579 individuals. We report on three main findings: (1) multivariate, data-driven statistics revealed a latent dimension of interindividual variation that linked advanced biological brain aging to lower cognitive and motor performance independent of chronological age, sex and education; (2) Vascular risk factors were significantly linked to PLS-derived aggregate measures of both biological brain aging as well as cognitive and motor performance; (3) Biological brain aging mediated the link between the vascular risk factors and cognitive as well as motor performance. These results applied to both investigated subcohorts: the UKB (discovery cohort) and the HCHS (replication cohort). In essence, our study demonstrates a clear link between biological brain aging and cognitive as well as motor abilities which also mediates the association between common vascular risk factors and impaired cognition and motor functioning.

### 4.1 PLS reveals a latent dimension integrating biological brain aging, cognition and motor function

We applied multivariate, data-driven statistics in form of a PLS in two large-scale population-based studies to map associative effects of three different global imaging markers of biological brain aging, i.e., relative brain age, WMH load and PSMD, to multiple measures of cognitive and motor function. The PLS models an underlying latent relationship that represents a multivariate mapping between imaging and clinical markers. The analysis identified a dominant latent variable associating higher markers biological brain aging with lower cognitive and motor scores, suggesting a direct link between increased biological brain aging and reduced performance in clinical tests. This variable accounted for 94.7% of the shared variance between imaging and clinical data, indicating a single axis of variation for the interplay between biological brain aging, cognitive and physical function. Bootstrapping of PLS results revealed that all biological brain aging markers as well as all clinical markers were significantly contributing to this association. Taken together, we interpret these findings as evidence that relative brain age, WMH and PSMD are considerably similar in their relationship to cognitive and motor performance and vice versa, i.e., the relationship is low dimensional. This notion is plausible given previous work showing high covariance of the investigated imaging markers as well as covariance between cognitive and motor test performances [11,37,38].

### 4.2 Biological brain aging is associated to worse cognitive performance across domains

Previous studies have individually connected single imaging markers of biological brain aging such as brain age gap, WMH, or PSMD with cognitive performance, yet the involvement of all cognitive domains is uncertain due to inconsistent findings [16,39,40]. Our results from PLS and multiple linear regression underscore that associations between brain aging and cognition are not confined to specific cognitive domains, a result likely attributable to our study’s ability to robustly detect subtle effects due to a large sample size. This aligns with previous research showing widespread influence of WMH and brain age gap measures across key cognitive areas [15,41]. An alternative hypothesis could be that the observed covariance in domain-specific cognitive tests stems from their shared reliance on certain cognitive functions, such as memory tests also tapping into attention and executive function.

Importantly, while all cognitive domains were found to be associated with brain aging, our findings highlight executive function and processing speed as the most strongly affected areas, consistent with existing literature linking age-related brain changes to declines in these faculties [5,42]. Furthermore, these results corroborate past studies connecting WMH and PSMD to specific impairments in executive function and processing speed, characteristic of cerebral small vessel disease [40,43].

### 4.3 Imaging markers of brain aging link to motor performance

Turning to motor function, we could show that imaging markers of biological brain aging link to motor performance, notably hand grip strength and physical activity measured via accelerometry, though the correlation was weaker than with cognitive function. The association between biological brain aging markers and motor function is less documented than that with cognitive measures; however, this link is supported by the known relationship between motor skills and cognition in mid to later life indicating shared variance [44]. Our findings align with previous studies demonstrating a negative correlation between brain aging and motor performance, specifically with hand grip strength and physical activity [45,46]. Pathomechanistically, older adults engage a more widespread network of brain areas for motor control, particularly the prefrontal cortex and basal ganglia, which are highly susceptible to aging [47]. This could lead to a mismatch in neural resource allocation. Moreover, decreased physical activity may both result from and contribute to biological brain aging. Given the neuroprotective benefits of physical exercise, a lack of it might increase the risk for onset and progression of neurodegenerative processes, highlighting an opportunity for targeted interventions [48].

### 4.4 Biological brain aging mediates the link between vascular risk, cognition and motor function

Vascular risk is a key modifiable factor influencing brain structure and function, with higher risk linked to poorer structural brain integrity and cognitive performance [18,49]. We examined the role of biological brain aging in mediating the relationship between vascular risk factors and cognitive/motor performance in a mediation analysis. Our findings show significant links between vascular risk factors (such as hypertension, dyslipidemia, glycemia, obesity, and smoking) and clinical PLS scores, which represent the overall adherence to the identified clinical covariance pattern - i.e., worse performance. This association was statistically mediated by the imaging PLS score, which captures the degree of biological brain aging. This implies that the association between vascular risk and cognitive as well as motor abilities is contingent on variations in biological brain aging markers, highlighting the importance of macrostructural and microstructural brain changes in clinical sequelae of age-related cerebrovascular disease [18,50]. Our results elucidate the connections between vascular risk and both structural and functional brain health, suggesting clinical applications. Modifying vascular risk through prevention and treatment could potentially mitigate biological brain aging. Future approaches could involve using brain imaging to tailor therapies and identify individuals most likely to benefit from interventions aimed at reducing cognitive and motor decline.

### 4.5 Strengths and limitations

Strengths of this work lie in its large sample size, which minimizes the overestimation of effects and enhances reproducibility [17]. Further strengths include the replication of findings in an independent sample, advanced neuroimaging and statistical techniques, and comprehensive cognitive phenotyping. However, there are limitations to this study. These include the cross-sectional study design, which limits causal interpretability. Longitudinal data would provide more definitive insights into these associations. Additionally, while the effect sizes observed were statistically significant, they were modest. It is important to note that these effect sizes were calculated after adjusting for chronological age, a major influencer of variance in cognitive and motor functions in population-based studies. Chronological age did not significantly influence the observed associative patterns, indicating that our analysis effectively isolated the impact of biological brain aging markers on motor and cognitive performance.

### 4.6 Conclusion

Drawing upon a comprehensive neuroimaging analysis, our research converges on a dominant latent dimension of interindividual differences that links biological brain aging to cognitive and motor performance. Our findings highlight the role of vascular risk factors in contributing to accelerated brain aging and worse cognitive and motor performance, advocating for the implementation of effective preventive strategies for upholding functional independence up until higher age.

## Acknowledgements

The authors wish to acknowledge all participants and staff of the UKB. This research has been conducted using the UKB Resource under Application Number 41655. This work uses data provided by patients and collected by the NHS as part of their care and support. Copyright © 2024, NHS England. Re-used with the permission of the NHS England and/or UKB. All rights reserved. Furthermore, this research used data assets made available by National Safe Haven as part of the Data and Connectivity National Core Study, led by Health Data Research UK in partnership with the Office for National Statistics and funded by UK Research and Innovation. Furthermore, we want to acknowledge all participants of the Hamburg City Health Study and cooperation partners, patrons and the Deanery from the University Medical Center Hamburg—Eppendorf for supporting the Hamburg City Health Study. Special thanks applies to the staff at the Epidemiological Study Center for conducting the study. The participating institutes and departments from the University Medical Center Hamburg-Eppendorf contribute all with individual and scaled budgets to the overall funding. The Hamburg City Health Study is also supported by Amgen, Astra Zeneca, Bayer, BASF, Deutsche Gesetzliche Unfallversicherung (DGUV), Deutsches Institut für Ernährungsforschung, the Innovative medicine initiative (IMI) under grant number No. 116074 and the Fondation Leducq under grant number 16 CVD 03., Novartis, Pfizer, Schiller, Siemens, Unilever and “Förderverein zur Förderung der HCHS e.V.”. The publication has been approved by the Steering Board of the Hamburg City Health Study.

## 7 Data availability

UKB data can be obtained via its standardized data access procedure (https://www.ukbiobank.ac.uk/). HCHS data can be obtained by qualified researcher on reasonable request to the study’s steering committee.

## 8 Author contributions

Each author has made a significant contribution to the manuscript and all authors read and approved its final version. We describe individual contributions to the paper using the CrediT contributor role taxonomy. Conceptualization: M.P., B.C.; Data curation: M.P., M.A.L., C.M., F.L.N., M.S., F.H.; Formal analysis: M.P., M.A.L.; Funding acquisition: S.B.E., G.T., B.C.; Investigation: M.P., M.A.L.; Methodology: M.P., M.A.L.; Software: M.P., M.A.L.; Supervision: G.T., B.C.; Visualization: M.P., M.A.L.; Writing—original draft: M.P., M.A.L.; Writing—review & editing: M.P., M.A.L., C.M., F.L.N., M.S., J.F., J.G., S.K., R.T., A.O., F.H., K.P., S.B.E., G.T., B.C.

## References

[1] Crimmins EM. Lifespan and Healthspan: Past, Present, and Promise. Gerontologist 2015;55:901–11. 10.1093/geront/gnv130.

[2] Krivanek TJ, Gale SA, McFeeley BM, Nicastri CM, Daffner KR. Promoting Successful Cognitive Aging: A Ten-Year Update. J Alzheimers Dis 2021;81:871–920. 10.3233/JAD-201462.

[3] Nyberg L, Lövdén M, Riklund K, Lindenberger U, Bäckman L. Memory aging and brain maintenance. Trends Cogn Sci 2012;16:292–305. 10.1016/j.tics.2012.04.005.

[4] de Lange A-MG, Anatürk M, Suri S, Kaufmann T, Cole JH, Griffanti L, et al. Multimodal brain-age prediction and cardiovascular risk: The Whitehall II MRI sub-study. Neuroimage 2020;222:117292. 10.1016/j.neuroimage.2020.117292.

[5] Petersen M, Nägele FL, Mayer C, Schell M, Rimmele DL, Petersen E, et al. Brain network architecture constrains age-related cortical thinning. Neuroimage 2022;264:119721. 10.1016/j.neuroimage.2022.119721.

[6] Storsve AB, Fjell AM, Tamnes CK, Westlye LT, Overbye K, Aasland HW, et al. Differential Longitudinal Changes in Cortical Thickness, Surface Area and Volume across the Adult Life Span: Regions of Accelerating and Decelerating Change. Journal of Neuroscience 2014;34:8488–98. 10.1523/JNEUROSCI.0391-14.2014.

[7] Storsve AB, Fjell AM, Yendiki A, Walhovd KB. Longitudinal Changes in White Matter Tract Integrity across the Adult Lifespan and Its Relation to Cortical Thinning. PLOS ONE 2016;11:e0156770. 10.1371/journal.pone.0156770.

[8] Frey BM, Petersen M, Schlemm E, Mayer C, Hanning U, Engelke K, et al. White matter integrity and structural brain network topology in cerebral small vessel disease: The Hamburg city health study. Hum Brain Mapp 2021;42:1406–15. 10.1002/hbm.25301.

[9] Ning K, Zhao L, Matloff W, Sun F, Toga AW. Association of relative brain age with tobacco smoking, alcohol consumption, and genetic variants. Sci Rep 2020;10:10. 10.1038/s41598-019-56089-4.

[10] Franke K, Gaser C. Ten Years of BrainAGE as a Neuroimaging Biomarker of Brain Aging: What Insights Have We Gained? Front Neurol 2019;10:789. 10.3389/fneur.2019.00789.

[11] Hofmann SM, Beyer F, Lapuschkin S, Goltermann O, Loeffler M, Müller K-R, et al. Towards the interpretability of deep learning models for multi-modal neuroimaging: Finding structural changes of the ageing brain. NeuroImage 2022;261:119504. 10.1016/j.neuroimage.2022.119504.

[12] Frey BM, Petersen M, Mayer C, Schulz M, Cheng B, Thomalla G. Characterization of White Matter Hyperintensities in Large-Scale MRI-Studies. Front Neurol 2019;10. 10.3389/fneur.2019.00238.

[13] Beaudet G, Tsuchida A, Petit L, Tzourio C, Caspers S, Schreiber J, et al. Age-Related Changes of Peak Width Skeletonized Mean Diffusivity (PSMD) Across the Adult Lifespan: A Multi-Cohort Study. Frontiers in Psychiatry 2020;11. 10.3389/fpsyt.2020.00342.

[14] Richard G, Kolskår K, Sanders A-M, Kaufmann T, Petersen A, Doan NT, et al. Assessing distinct patterns of cognitive aging using tissue-specific brain age prediction based on diffusion tensor imaging and brain morphometry. PeerJ 2018;6:e5908. 10.7717/peerj.5908.

[15] Jawinski P, Markett S, Drewelies J, Düzel S, Demuth I, Steinhagen-Thiessen E, et al. Linking Brain Age Gap to Mental and Physical Health in the Berlin Aging Study II. Frontiers in Aging Neuroscience 2022;14.

[16] Boyle R, Jollans L, Rueda-Delgado LM, Rizzo R, Yener GG, McMorrow JP, et al. Brain-predicted age difference score is related to specific cognitive functions: a multi-site replication analysis. Brain Imaging and Behavior 2021;15:327–45. 10.1007/s11682-020-00260-3.

[17] Marek S, Tervo-Clemmens B, Calabro FJ, Montez DF, Kay BP, Hatoum AS, et al. Reproducible brain-wide association studies require thousands of individuals. Nature 2022;603:654–60. 10.1038/s41586-022-04492-9.

[18] Petersen M, Hoffstaedter F, Nägele FL, Mayer C, Schell M, Rimmele DL, et al. A latent clinical-anatomical dimension relating metabolic syndrome to brain structure and cognition. eLife 2024;12:RP93246. 10.7554/eLife.93246.

[19] Petersen M, Frey BM, Schlemm E, Mayer C, Hanning U, Engelke K, et al. Network Localisation of White Matter Damage in Cerebral Small Vessel Disease. Sci Rep 2020;10:9210. 10.1038/s41598-020-66013-w.

[20] Fawns-Ritchie C, Deary IJ. Reliability and validity of the UK Biobank cognitive tests. PLoS One 2020;15:e0231627. 10.1371/journal.pone.0231627.

[21] Coenen M, Kuijf HJ, Huenges Wajer IMC, Duering M, Wolters FJ, Fletcher EF, et al. Strategic white matter hyperintensity locations for cognitive impairment: A multicenter lesion-symptom mapping study in 3525 memory clinic patients. Alzheimer’s & Dementia 2023;19:2420–32. 10.1002/alz.12827.

[22] Nevill AM, Tomkinson GR, Lang JJ, Wutz W, Myers TD. How Should Adult Handgrip Strength Be Normalized? Allometry Reveals New Insights and Associated Reference Curves. Med Sci Sports Exerc 2022;54:162–8. 10.1249/MSS.0000000000002771.

[23] Fischl B, Dale AM. Measuring the thickness of the human cerebral cortex from magnetic resonance images. Proc Natl Acad Sci U S A 2000;97:11050–5. 10.1073/pnas.200033797.

[24] Dale AM, Fischl B, Sereno MI. Cortical surface-based analysis. I. Segmentation and surface reconstruction. Neuroimage 1999;9:179–94. 10.1006/nimg.1998.0395.

[25] Fischl B, Salat DH, Busa E, Albert M, Dieterich M, Haselgrove C, et al. Whole brain segmentation: automated labeling of neuroanatomical structures in the human brain. Neuron 2002;33:341–55. 10.1016/s0896-6273(02)00569-x.

[26] Fischl B, Dale AM. Measuring the thickness of the human cerebral cortex from magnetic resonance images. Proceedings of the National Academy of Sciences 2000;97:11050–5. 10.1073/pnas.200033797.

[27] Desikan RS, Ségonne F, Fischl B, Quinn BT, Dickerson BC, Blacker D, et al. An automated labeling system for subdividing the human cerebral cortex on MRI scans into gyral based regions of interest. Neuroimage 2006;31:968–80. 10.1016/j.neuroimage.2006.01.021.

[28] Smith SM, Vidaurre D, Alfaro-Almagro F, Nichols TE, Miller KL. Estimation of brain age delta from brain imaging. Neuroimage 2019;200:528–39. 10.1016/j.neuroimage.2019.06.017.

[29] Bretzner M, Bonkhoff AK, Schirmer MD, Hong S, Dalca A, Donahue K, et al. Radiomics-Derived Brain Age Predicts Functional Outcome After Acute Ischemic Stroke. Neurology 2022:10.1212/WNL.0000000000201596. 10.1212/WNL.0000000000201596.

[30] Hamdan S, More S, Sasse L, Komeyer V, Patil KR, Raimondo F, et al. Julearn: an easy-to-use library for leakage-free evaluation and inspection of ML models. Gigabyte 2024;2024:1–16. 10.46471/gigabyte.113.

[31] Sanford N, Ge R, Antoniades M, Modabbernia A, Haas SS, Whalley HC, et al. Sex differences in predictors and regional patterns of brain age gap estimates. Human Brain Mapping 2022;43:4689–98. 10.1002/hbm.25983.

[32] Griffanti L, Zamboni G, Khan A, Li L, Bonifacio G, Sundaresan V, et al. BIANCA (Brain Intensity AbNormality Classification Algorithm): A new tool for automated segmentation of white matter hyperintensities. Neuroimage 2016;141:191–205. 10.1016/j.neuroimage.2016.07.018.

[33] Petersen M, Nägele FL, Mayer C, Schell M, Petersen E, Kühn S, et al. Brain imaging and neuropsychological assessment of individuals recovered from mild to moderate SARS-CoV-2 infection 2022:2022.07.08.22277420. 10.1101/2022.07.08.22277420.

[34] Zanon Zotin MC, Yilmaz P, Sveikata L, Schoemaker D, van Veluw SJ, Etherton MR, et al. Peak Width of Skeletonized Mean Diffusivity: A Neuroimaging Marker for White Matter Injury. Radiology 2023:212780. 10.1148/radiol.212780.

[35] Smith SM, Jenkinson M, Johansen-Berg H, Rueckert D, Nichols TE, Mackay CE, et al. Tract-based spatial statistics: voxelwise analysis of multi-subject diffusion data. Neuroimage 2006;31:1487–505. 10.1016/j.neuroimage.2006.02.024.

[36] Baron RM, Kenny DA. The moderator-mediator variable distinction in social psychological research: conceptual, strategic, and statistical considerations. J Pers Soc Psychol 1986;51:1173–82. 10.1037//0022-3514.51.6.1173.

[37] Busby N, Newman-Norlund S, Sayers S, Newman-Norlund R, Wilson S, Nemati S, et al. White matter hyperintensity load is associated with premature brain aging. Aging (Albany NY) 2022;14:9458–65. 10.18632/aging.204397.

[38] Lee P-L, Kuo C-Y, Wang P-N, Chen L-K, Lin C-P, Chou K-H, et al. Regional rather than global brain age mediates cognitive function in cerebral small vessel disease. Brain Communications 2022;4:fcac233. 10.1093/braincomms/fcac233.

[39] Prins ND, Scheltens P. White matter hyperintensities, cognitive impairment and dementia: an update. Nat Rev Neurol 2015;11:157–65. 10.1038/nrneurol.2015.10.

[40] Deary IJ, Ritchie SJ, Muñoz Maniega S, Cox SR, Valdés Hernández MC, Luciano M, et al. Brain Peak Width of Skeletonized Mean Diffusivity (PSMD) and Cognitive Function in Later Life. Front Psychiatry 2019;10:524. 10.3389/fpsyt.2019.00524.

[41] Hamilton OKL, Backhouse EV, Janssen E, Jochems ACC, Maher C, Ritakari TE, et al. Cognitive impairment in sporadic cerebral small vessel disease: A systematic review and meta-analysis. Alzheimer’s & Dementia 2021;17:665–85. 10.1002/alz.12221.

[42] Heckner MK, Cieslik EC, Eickhoff SB, Camilleri JA, Hoffstaedter F, Langner R. The Aging Brain and Executive Functions Revisited: Implications from Meta-analytic and Functional-Connectivity Evidence. J Cogn Neurosci 2021;33:1716–52. 10.1162/jocn_a_01616.

[43] Low A, Mak E, Stefaniak JD, Malpetti M, Nicastro N, Savulich G, et al. Peak Width of Skeletonized Mean Diffusivity as a Marker of Diffuse Cerebrovascular Damage. Frontiers in Neuroscience 2020;14.

[44] Duchowny KA, Ackley SF, Brenowitz WD, Wang J, Zimmerman SC, Caunca MR, et al. Associations Between Handgrip Strength and Dementia Risk, Cognition, and Neuroimaging Outcomes in the UK Biobank Cohort Study. JAMA Network Open 2022;5:e2218314. 10.1001/jamanetworkopen.2022.18314.

[45] Fleischman DA, Yang J, Arfanakis K, Arvanitakis Z, Leurgans SE, Turner AD, et al. Physical activity, motor function, and white matter hyperintensity burden in healthy older adults. Neurology 2015;84:1294–300. 10.1212/WNL.0000000000001417.

[46] Cole JH, Ritchie SJ, Bastin ME, Valdés Hernández MC, Muñoz Maniega S, Royle N, et al. Brain age predicts mortality. Mol Psychiatry 2018;23:1385–92. 10.1038/mp.2017.62.

[47] Raz N, Gunning FM, Head D, Dupuis JH, McQuain J, Briggs SD, et al. Selective aging of the human cerebral cortex observed in vivo: differential vulnerability of the prefrontal gray matter. Cereb Cortex 1997;7:268–82. 10.1093/cercor/7.3.268.

[48] Ahlskog JE, Geda YE, Graff-Radford NR, Petersen RC. Physical Exercise as a Preventive or Disease-Modifying Treatment of Dementia and Brain Aging. Mayo Clinic Proceedings 2011;86:876. 10.4065/mcp.2011.0252.

[49] Borshchev YYu, Uspensky YP, Galagudza MM. Pathogenetic pathways of cognitive dysfunction and dementia in metabolic syndrome. Life Sciences 2019;237:116932. 10.1016/j.lfs.2019.116932.

[50] Petersen M, Chevalier C, Naegele FL, Ingwersen T, Omidvarnia A, Hoffstaedter F, et al. Mapping the interplay of atrial fibrillation, brain structure, and cognitive dysfunction. Alzheimer’s & Dementia 2024;20:4512–26. 10.1002/alz.13870.

