## Supplementals for "Markers of Biological Brain Aging Mediate Effects of Vascular Risk Factors on Cognitive and Motor Functions: A Multivariate Imaging Analysis of 40,579 Individuals"

### Methods

#### Text S1 – Description of motor and cognitive tests

**Motor tests – UKB**

**Hand grip strength**

Hand grip strength assessment was performed using a Jamar J00105 hydraulic hand dynamometer. Holding the dynamometer, the participant's arm was positioned against their torso, elbow at 90° and thumb facing upwards. Grip force assessment lasted for 3 seconds and is measured in kilograms with a peak-hold needle that remains in place once grip was released. Measurements were performed one time for each hand. (https://biobank.ndph.ox.ac.uk/showcase/refer.cgi?id=100232)

**Average acceleration**

Physical activity was measured using the Axivity AX3 wrist-worn triaxial accelerometer, a commercial version of the Open Movement AX3 open source sensor (https://github.com/digitalinteraction/openmovement) designed by Open Lab, Newcastle University. Wrist accelerometers were shipped via post, worn by participants for one week and then send back via post. Non-wearing data episodes were imputed using the average of similar time of day data to create individual accelerometry profiles.[1] (https://biobank.ndph.ox.ac.uk/showcase/refer.cgi?id=169649)

**Cognitive tests – UKB**

The cognitive test battery of the UKB represent computerized adaptions of the original tests.[2]

**Numeric Memory Test (max digit)**

The Numeric Memory Test assesses short-term memory functioning. A two-digit number was presented to the participant followed by a recall test for the number. For each correct recall test a consequent digit was added and the procedure was repeated. The number was extended up to a twelve-digit number. Number of rounds were documented and used as evaluation metric. (https://biobank.ndph.ox.ac.uk/showcase/refer.cgi?id=6)

**Trail Making Test A and B (seconds)**

The Trail Making Test A and B evaluate processing speed and executive functioning. In part A, the participant was asked to draw lines sequentially connecting 25 encircled numbers. In part B, the participant had to alternate between sequentially

increasing numbers and lexicographically increasing letters. Test performance was measured in time (deciseconds) that is needed to finish the task.
(https://biobank.ndph.ox.ac.uk/showcase/refer.cgi?id=8481, https://biobank.ndph.ox.ac.uk/showcase/refer.cgi?id=105)

**Matrix Pattern Completion Test (correct puzzles)**
The Matrix Pattern Completion Test evaluates non-verbal fluid reasoning ability. The test was performed on a computer. A puzzle pattern was presented to the participant and the participant was asked to complete the pattern by selecting from a range of pattern pieces the one that best complements the presented pattern. After a correct pattern completion, a new pattern was shown. The maximal number of displayed patterns was 15. The number of successful pattern completions was used as scoring metric.
(https://biobank.ndph.ox.ac.uk/showcase/label.cgi?id=501, https://biobank.ndph.ox.ac.uk/showcase/refer.cgi?id=15)

**Fluid intelligence**
The fluid intelligence examination tests for logic and reasoning ability, independent of acquired knowledge. The participant was asked to answer as many questions as possible within two minutes from a questionnaire with 13 questions overall. Skipping of questions was possible. The number of correctly answered questions represented the scoring metric.
(https://biobank.ndph.ox.ac.uk/showcase/refer.cgi?id=100231)

**Reaction time**
Reaction time was measured by 12 rounds of the card game „Snap“. Two cards displaying a pattern were presented on a screen. In case of matching cards, the participant was asked to press a button as fast as possible. Scores were measured as the mean reaction time in milliseconds for correct answers.
(https://biobank.ndph.ox.ac.uk/showcase/refer.cgi?id=100245)

**Paired Associate Learning Test (correct pairs)**
The Paired Associate Learning Test examines verbal declarative memory. 12 word pairs were presented on a screen to the participant. After 30 seconds of memorizing, the participant continued with another test. After that a list of ten single words without their match were presented to the participant. The participant was asked to find the corresponding word pair from a choice of four words displayed on the screen. Absolute number of correct word pairs represented the scoring metric.
(https://biobank.ndph.ox.ac.uk/showcase/refer.cgi?id=2561)

**Tower Rearranging Test (Correct puzzles)**
The Tower Rearranging Test evaluates executive function with an emphasis on planning. An image displaying three pegs and three differently coloured hops was

presented to the participant. In the following, 18 images with different hop positions were shown and for each image the participant was asked to indicate how many moves were needed to rearrange the hops so that their position match again. The absolute number of correctly solved images was used as scoring metric.
(https://biobank.ndph.ox.ac.uk/showcase/label.cgi?id=503,
https://biobank.ndph.ox.ac.uk/showcase/refer.cgi?id=527)

**Symbol Digit Substitution Test (correct matches)**
The Symbol Digit Substitution Test examines complex processing speed. A table listing eight numbers and their corresponding symbol was presented to the participant. The participant was then asked to translate an eight-symbol combination into their corresponding number combination as fast as possible. The test lasted for one minute. Absolute number of correctly translated symbol-number combinations was used as scoring metric.
(https://biobank.ndph.ox.ac.uk/showcase/refer.cgi?id=2103)

#### Motor tests – HCHS

**Hand grip strength**
Hand grip strength assessment was performed using a Jamar Plus Digital Dynamometer (Patterson Medical, Sammons Preston, Bolingbrook, IL). Holding the dynamometer the participant's arm was positioned against their torso, elbow at 90° and thumb facing upwards. Grip force assessment lasted for 3 seconds and is measured in kilograms. Each hand was measured three times with 15 seconds in between each measurement. The resulting measurements were averaged for each hand.

**Timed Up And Go Test**
The Timed Up And Go Test measures basic physical mobility and the risk of falling [3]. The test consisted of the participant standing up from a seated position, walking three meters, turning around and returning to the chair to sit down again. Time in seconds was measured and used as scoring metric. Time scores of <10s, 10-19s, 20-29s, >30s respectively represent no, no functionally relevant, functionally relevant and distinctively, functionally relevant mobility restriction.

#### Cognitive tests – HCHS

In the HCHS cohort all baseline evaluations, standardized neuropsychological tests and motor assessments were performed by specifically trained medical professionals.

**International Standard Classification of Education questionnaire (ISCED)**

In HCHS the ISCED version from 2011 was used. Furthermore, subject ISCED scores were subdivided into categories low (0-1), medium (3-4) and high education (5-8).

**Animal Naming Test**

The Animal Naming Test assesses verbal fluency. Participants were asked to name as many animal names as possible within 60 seconds. Absolute number of animals was counted and used as test scoring metric.[4]

**Trail Making Test A and B**

The Trail Making Test A and B evaluate processing speed and executive functioning. In part A, the participant was asked to draw lines sequentially connecting 25 encircled numbers that were spread across a sheet of paper. In part B the participant had to alternate between sequentially increasing numbers and lexicographically increasing letters. Test performance was measured in seconds that is needed to finish the task.[5]

**Multiple Choice Vocabulary Intelligence Test B**

The Vocabulary Test B evaluates general intelligence levels. The test consists of 40 items of five words each. A selection of five words was presented to the participant. One word had a meaning, the other four were fictional. The participant was asked to identify the word with meaning. The number of correctly identified words served as scoring metric.[6]

**Word List Recall Test**

The Word Recall Test examines verbal learning. A word list of ten common nouns was presented and read out loud by a test participant. Subsequently an immediate recall test was performed. Test scores were collected for the recall test (0-10 points), giving one point for each remembered word.[6]

### Text S2: MRI acquisition and preprocessing

#### MRI acquisition

The full UKB neuroimaging protocol is available online (https://biobank.ctsu.ox.ac.uk/crystal/crystal/docs/brain_mri.pdf).[7] MR images were acquired on a 3-T Siemens Skyra MRI scanner 3-T (Siemens, Erlangen, Germany) with the following acquisition parameters. 3D T1-weighted MRI - 3D MPRAGE sequence – TR = 2000 ms, TE = 2.01 ms, 256 axial slices, slice thickness = 1 mm, and in-plane resolution = 1×1 mm. Fluid attenuated inversion recovery MRI was based on the following parameters. Resolution = 1.05×1×1 mm; field-of-view = 192×256×256 matrix, TI/TR = 1800/5000 ms, fat saturation. Multi-shell diffusion-weighted imaging was based on the following sequence parameters: 2×2×2 mm, field-of-view 104×104×72, TR = 3600 ms, TE = 92 ms, 50 diffusion-encoding directions — 50x b = 1000 s/mm$^2$ and 50x b = 2000 s/mm$^2$.

HCHS MRI image acquisition was conducted on a single 3-T Siemens Skyra MRI scanner (Siemens, Erlangen, Germany). The acquisition protocol was described previously.[8] In brief, images were acquired using the following acquisition parameters: 3D T1-weighted MRI – 3D MPRAGE sequence – TR = 2500 ms, TE = 2.12 ms, 256 axial slices, slice thickness = 0.94 mm, and IPR = 0.83×0.83 mm; FLAIR: TR = 4700 ms, TE = 392 ms, 192 axial slices, slice thickness = 0.9 mm, and in-plane resolution = 0.75×0.75 mm; Single-shell diffusion weighted imaging: TR = 8500 ms, TE = 75 ms, 75 axial slices, slice thickness = 2 mm, in-plane resolution = 2×2mm, 64 noncollinear gradient directions with b = 1000 s/mm$^2$, 1 image with b = 0 s/mm).[9]

#### MRI preprocessing

UKB MR image preprocessing protocols for T1 and DWI images are documented online (https://biobank.ctsu.ox.ac.uk/crystal/crystal/docs/brain_mri.pdf).

In the HCHS, T1 weighted MRI images were further preprocessed using fMRIPrep 20.2.6. Image preprocessing included correction for intensity non-uniformity (INU) with N4BiasFieldCorrection,[10] distributed with ANTs 2.3.3 (https://github.com/ANTsX/ANTs). Data outputs were used as T1w-reference throughout the ongoing workflow. The T1w-reference was then skull-stripped with a Nipype implementation of the antsBrainExtraction.sh workflow (from ANTs), using OASIS30ANTs as target template. The preprocessed T1w images were propagated to the canonical FreeSurfer pipeline (v. 6.0.1) for subcortical segmentation and surface reconstruction.[11–13]

QSIPrep 0.14.2[14] was used for preprocessing of diffusion-weighted MRI (dMRI). MP-PCA denoising as implemented in MRtrix3's dwidenoise[15] was applied with a 5-voxel window. After 3 MP-PCA, Gibbs unringing was performed using MRtrix3's

mrdegibbs.[16] Following unringing, B1 field inhomogeneity was corrected using dwibiascorrect from MRtrix3 with the N4 algorithm.[10]
FSL (version 6.0.3:b862cdd5)'s eddy was used for head motion correction and eddy current correction.[17] Eddy was configured with a $q$-space smoothing factor of 10, a total of 5 iterations, and 1000 voxels used to estimate hyperparameters. A linear first level model and a linear second level model were used to characterize eddy current-related spatial distortion. $q$-space coordinates were forcefully assigned to shells. Field offset was attempted to be separated from subject movement. Shells were aligned post-eddy. Eddy's outlier replacement was run. Data were grouped by slice, only including values from slices determined to contain at least 250 intracerebral voxels. Groups deviating by more than 4 standard deviations from the prediction had their data replaced with imputed values. Final interpolation was performed using the jac method.
A deformation field to correct for susceptibility distortions was estimated based on fMRI-prep's fieldmap-less approach.[18] The deformation field results from co-registering the b0 reference to the same-subject T1w-reference with its intensity inverted.[19] Registration was performed with antsRegistration (ANTs 2.3.1), and the process regularized by constraining deformation to be nonzero only along the phase-encoding direction and modulated with an average fieldmap template. Based on the estimated susceptibility distortion, an unwarped b=0 reference was calculated for a more accurate co-registration with the anatomical reference. Several confounding time-series were calculated based on the preprocessed DWI: framewise displacement using the implementation in Nipype (following the definitions by [20]). The head motion estimates calculated in the correction step were also placed within the corresponding confounds file. Slicewise cross correlation was also calculated. The DWI time-series were resampled to ACPC, generating a preprocessed DWI run in ACPC space with 2mm isotropic voxels.

Many internal operations of QSIPrep use Nilearn[21] and Dipy.[22] For more details of the pipeline, see the section corresponding to workflows in QSIPrep's documentation.[23,24]

##### Freesurfer

In the UKB, cortical surface reconstruction and subcortical gray and white matter volumetry were performed based on FreeSurfer version 6.0 (version=6-20170118 build-stamp=v6.0.0-2beb96c). After segmentation, FreeSurfer outputs were quality checked based on the Qoala-T approach.[25] For the presented analysis, regional measurements of cortical thickness and subcortical volumes precomputed by the UKB were used.

In the HCHS, preprocessed T1 weighted MRI images served as input data for FreeSurfer v.7.1. Reconstructions of cortical surfaces and volumetric segmentations of subcortical gray and white matter were performed. After surface reconstruction, surface parcellation was performed using the Desikan-Killany cortical atlas. The assessment of subcortical white and gray matter was conducted via nonlinear registration into the talairach space.[26–28] DWI images were processed employing QSIPrep 0.14.21.[14] For more detailed insights into the used T1 weighted and DWI image preprocessing pipelines please refer to supplements (Supplement 2).[26] Image quality control is described elsewhere (https://github.com/csi-hamburg/hchs_qa/wiki/Quality-Assessment-of-Neuroimaging-Data-of-the-Hamburg-City-Health-Study).

For the following analysis, cortical thickness and subcortical volumes of 99 regions of interest were used: lh_bankssts_thickness, lh_caudalanteriorcingulate_thickness, lh_caudalmiddlefrontal_thickness, lh_cuneus_thickness, lh_entorhinal_thickness, lh_fusiform_thickness, lh_inferiorparietal_thickness, lh_inferiortemporal_thickness, lh_isthmuscingulate_thickness, lh_lateraloccipital_thickness, lh_lateralorbitofrontal_thickness, lh_lingual_thickness, lh_medialorbitofrontal_thickness, lh_middletemporal_thickness, lh_parahippocampal_thickness, lh_paracentral_thickness, lh_parsopercularis_thickness, lh_parsorbitalis_thickness, lh_parstriangularis_thickness, lh_pericalcarine_thickness, lh_postcentral_thickness, lh_posteriorcingulate_thickness, lh_precentral_thickness, lh_precuneus_thickness, lh_rostralanteriorcingulate_thickness, lh_rostralmiddlefrontal_thickness, lh_superiorfrontal_thickness, lh_superiorparietal_thickness, lh_superiortemporal_thickness, lh_supramarginal_thickness, lh_frontalpole_thickness, lh_transversetemporal_thickness, lh_insula_thickness, rh_bankssts_thickness, rh_caudalanteriorcingulate_thickness, rh_caudalmiddlefrontal_thickness, rh_cuneus_thickness, rh_entorhinal_thickness, rh_fusiform_thickness, rh_inferiorparietal_thickness, rh_inferiortemporal_thickness, rh_isthmuscingulate_thickness, rh_lateraloccipital_thickness, rh_lateralorbitofrontal_thickness, rh_lingual_thickness, rh_medialorbitofrontal_thickness, rh_middletemporal_thickness, rh_parahippocampal_thickness, rh_paracentral_thickness, rh_parsopercularis_thickness, rh_parsorbitalis_thickness, rh_parstriangularis_thickness, rh_pericalcarine_thickness, rh_postcentral_thickness, rh_posteriorcingulate_thickness, rh_precentral_thickness, rh_precuneus_thickness, rh_rostralanteriorcingulate_thickness, rh_rostralmiddlefrontal_thickness, rh_superiorfrontal_thickness, rh_superiorparietal_thickness, rh_superiortemporal_thickness, rh_supramarginal_thickness, rh_frontalpole_thickness, rh_transversetemporal_thickness, rh_insula_thickness, Left-Lateral-Ventricle, Left-Inf-Lat-Vent, Left-Cerebellum-White-Matter, Left-Cerebellum-Cortex, Left-Thalamus-Proper, Left-Caudate, Left-Putamen, Left-Pallidum, 3rd-Ventricle, 4th-Ventricle, Left-Hippocampus, Left-Amygdala, Left-Accumbens-area, Left-VentralDC, Left-choroid-plexus, Right-Lateral-Ventricle, Right-Inf-Lat-Vent, Right-Cerebellum-White-Matter, Right-Cerebellum-Cortex, Right-Thalamus-Proper, Right-Caudate, Right-Putamen, Right-Pallidum, Right-Hippocampus, Right-Amygdala, Right-Accumbens-area, Right-VentralDC, Rightchoroid-plexus, CC_Posterior, CC_Mid_Posterior, CC_Central, CC_Mid_Anterior, CC_Anterior.

**White matter hyperintensity (WMH) segmentation**

In the UKB, WMH Segmentation was based on FLAIR and T1w MR images. Lesion segmentation was automatically performed using the BIANCA tool from Griffanti et al..[29] WMH and intracranial volumes were directly downloaded from the UKB databank. WMHs loads were calculated as the ratio of WMH volume and intracranial volume.

As described previously, in the HCHS WMH segmentation was performed using FSL's Brain Intensity AbNormality Classification Algorithm (BIANCA) with LOCally Adaptive Threshold Estimation (LOCATE).[26] In short BIANCA input data consisted of (1) a brain-extracted FLAIR image, (2) a FLAIR image rigid-registration matrix to MNI space, (3) T1w image rigidly registered to FLAIR space and (4) manually segmented WMH masks as the gold standard. Manual WMH segmentation was performed by two expert raters (M.P., C.M.) resulting in a training dataset of 100 individuals. The training dataset was then used to train a supervised k-nearest neighbor algorithm (BIANCA). Raw output masks of BIANCA were used as input for LOCATE to enable region specific thresholding and thus optimizing segmentation precision. Additional input data for LOCATE included (1) the corresponding brain-extracted FLAIR, (2) in FLAIR space rigidly registered T1w images, (3) a ventricle distancemap, (4) the manual segmentations of training dataset in FLAIR and 5.) a brain mask in FLAIR. Ongoing FreeSurfer-derived masks of the dilated cortical ribbon, eroded ventricle ribbon, corpus callosum and basal ganglia mask were aggregated to exclude non-white matter tissue from the LOCATE outputs. WMH loads were calculated as the ratio of the WMH volume and the intracranial volume.[29,30]

### Table S3: Age prediction scores

##### UKB

| CV fold | $R^2$ – male | Neg. MAE – male | Neg. MSE – male | $R^2$ – female | Neg. MAE – female | Neg. MSE – female |
|---|---|---|---|---|---|---|
| 1 | 0.518 | -4.418 | -30.073 | 0.460 | -4.432 | -30.536 |
| 2 | 0.536 | -4.264 | -28.153 | 0.471 | -4.338 | -29.295 |
| 3 | 0.493 | -4.360 | -29.403 | 0.475 | -4.386 | -29.718 |
| 4 | 0.507 | -4.369 | -29.299 | 0.424 | -4.586 | -32.716 |
| 5 | 0.510 | -4.338 | -29.315 | 0.445 | -4.490 | -31.393 |
| Mean ± SD | 0.513 ± 0.014 | -4.350 ± 0.050 | -29.249 ± 0.618 | 0.454 ± 0.018 | -4.447 ± 0.086 | -30.732 ± 1.225 |

##### HCHS

| CV fold | $R^2$ – male | Neg. MAE – male | Neg. MSE – male | $R^2$ – female | Neg. MAE – female | Neg. MSE – female |
|---|---|---|---|---|---|---|
| 1 | 0.403 | -4.689 | -34.354 | 0.424 | -5.244 | -40.871 |
| 2 | 0.488 | -4.679 | -32.675 | 0.479 | -4.952 | -38.278 |
| 3 | 0.489 | -4.768 | -35.672 | 0.449 | -5.080 | -39.131 |
| 4 | 0.499 | -4.873 | -35.767 | 0.476 | -5.082 | -39.594 |
| 5 | 0.484 | -4.788 | -35.466 | 0.407 | -4.998 | -39.128 |
| Mean ± SD | 0.473 ± 0.035 | -4.759 ± 0.071 | -34.787 ± 1.171 | 0.447 ± 0.028 | -5.071 ± 0.099 | -39.401 ± 0.849 |

Scores representing prediction accuracy of models predicting chronological age from regional brain morphology. Listed metrics are $R^2$, mean absolute error (MAE) and mean squared error (MSE). The results for each accuracy metric are presented separately for each cross-validation fold and as means with standard deviations (SD).

#### Figure S4: Chronological age correlation of relative brain age

**UKB**

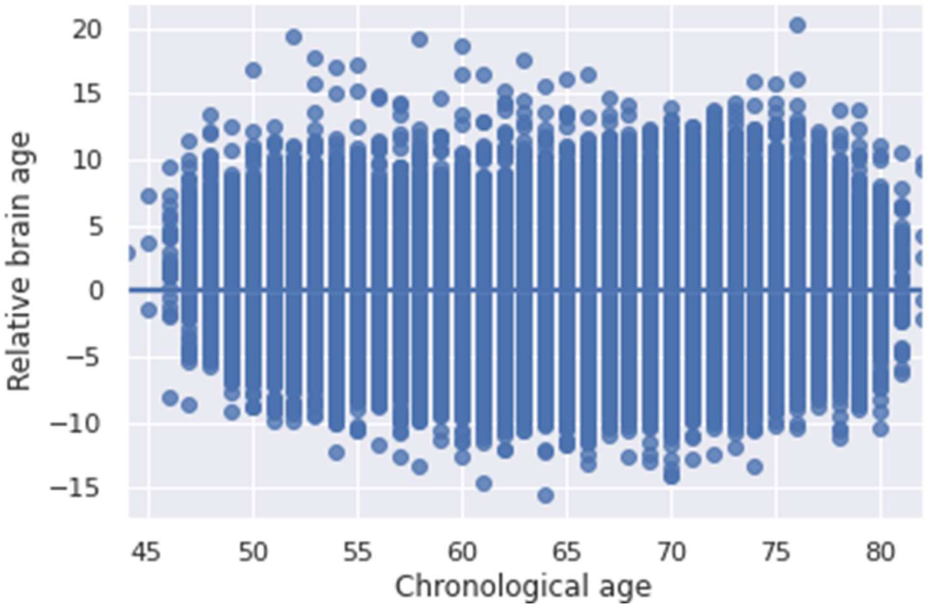

**HCHS**

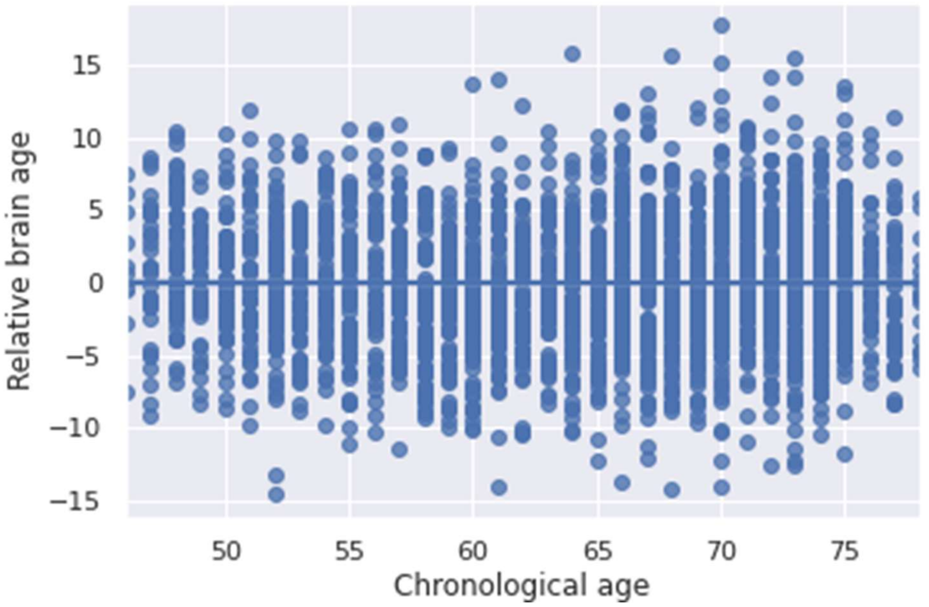

The regression plots display the linear relationship of relative brain age and chronological age. The variables are orthogonal indicating sufficient correction for chronological age during the computation of relative brain age.

### Figure S5: Age correlation of imaging and clinical variables before residualization - UKB

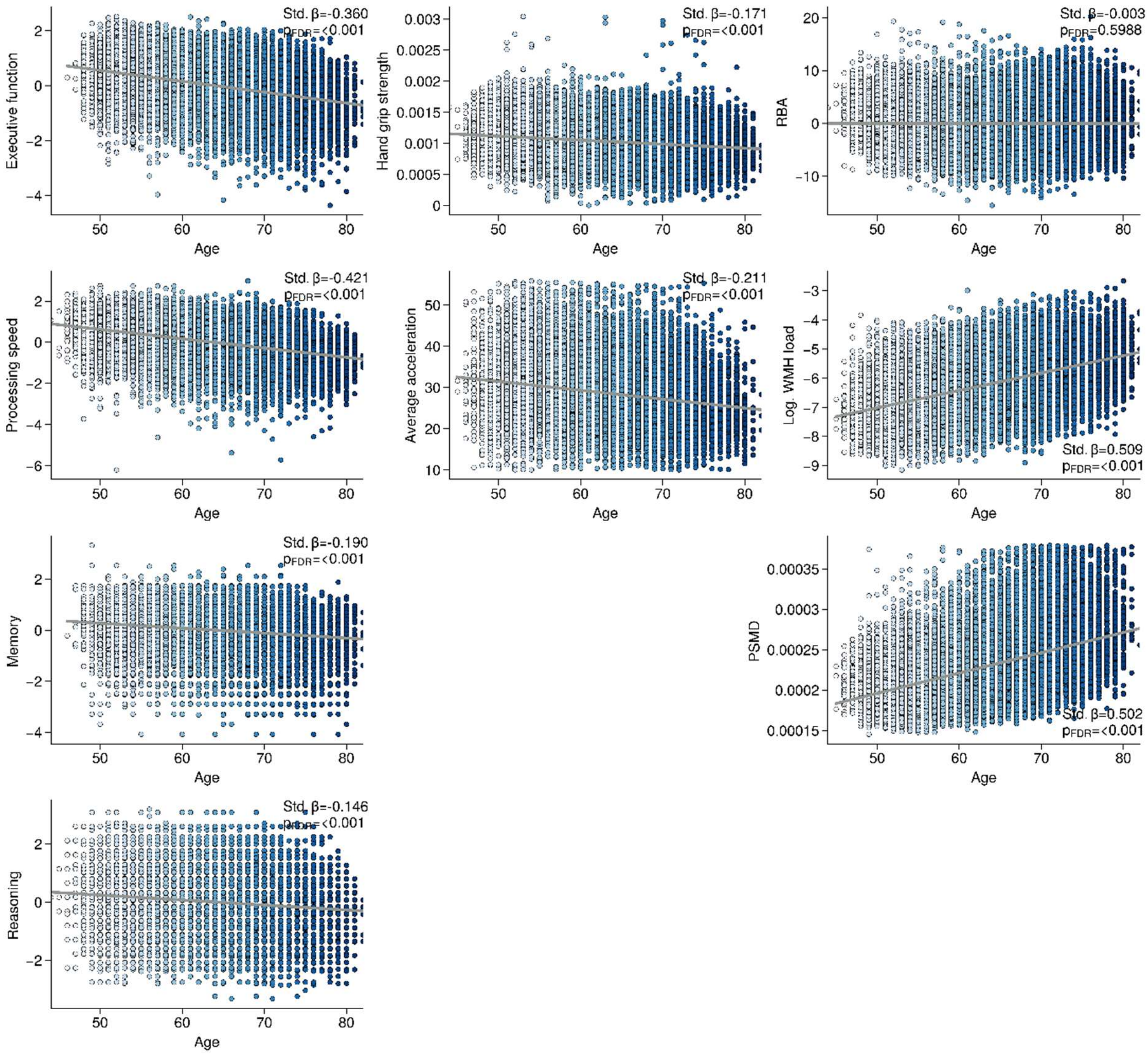

Regression plots indicate the relationships between chronological age to imaging and clinical markers before residualization (i.e., correction for age).

### Figure S6: Age correlation of imaging and clinical variables before residualization - HCHS

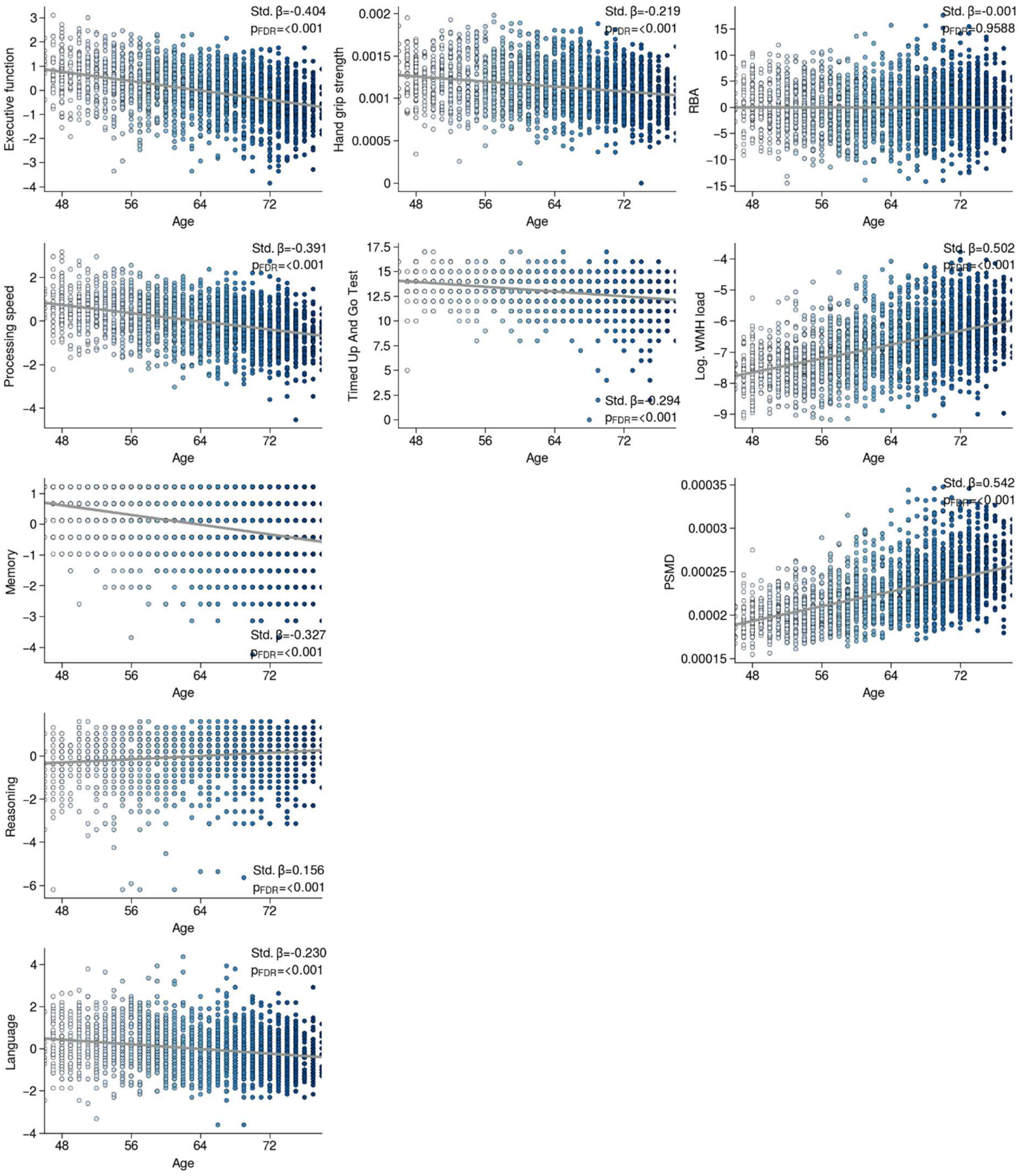

Regression plots indicate the relationships between chronological age to imaging and clinical markers before residualization (i.e., correction for age).

### Text S7: Methodological details of partial least squares correlation

In the current study, we employ PLS to examine the multivariate relationship between imaging markers of biological brain age and measures of cognitive and motor performance. After deconfounding for covariates (age, sex, education) two datasets $X_{n_{subjects} \times n_{imaging\ variables}}$ and $y_{n_{subjects} \times n_{clinical\ variables}}$ serve as input data for the PLS. In a first step, a cross-covariance matrix is calculated ($R_{n_{imaging\ variables} \times n_{clinical\ variables}}$). This correlation matrix is further propagated to singular value decomposition resulting in a set of latent variables. The number of latent variables is determined by the number of columns of the smaller initial input matrix. In case of our study, $X_{n_{subjects} \times n_{imaging\ variables}}$ with the number of n=3 imaging variables resulting in three latent variables. A latent variable is defined as a left singular vector $U_{n_{imaging\ variables} \times n_{imaging\ loading}}$ and a right singular vector $V_{n_{clinical\ variables} \times n_{clinical\ loadings}}$ and a singular value. The singular vectors encompass a loading for each original variable that if linearly combined maximizes the covariance of both variable sets. The explained covariance is calculated as the ratio of the squared singular value to the sum of all squared singular values. To test the significance of a latent variable, the empiric explained covariance value is compared to the distribution of permuted explained covariance values. This is obtained via permutation (n=5000) of the subject order in $X_{n_{subjects} \times n_{imaging\ variables}}$ (Fig. 1a). Subject-level imaging and clinical scores are calculated via the linear combination of $X_{n_{subjects} \times n_{imaging\ variables}}$ and $U_{n_{imaging\ variables} \times n_{imaging\ loadings}}$ as well as $y_{n_{subjects} \times n_{clinical\ variables}}$ and $V_{n_{clinical\ variables} \times n_{clinical\ loadings}}$, respectively. The higher the resulting subject-level imaging and clinical score, the stronger an individual expresses the imaging or clinical phenotype defined by the corresponding covariance pattern.

Bootstrapping is performed to measure the contribution of each imaging and clinical variable to the imaging-clinical relationship. Subsets of $X_{n_{subjects} \times n_{imaging\ variables}}$ and $y_{n_{subjects} \times n_{clinical\ variables}}$ are resampled (n=5000) with replacement, their correlation matrices are determined and further propagated to singular value decomposition. Bootstrapping yields variable-specific sampling distributions of singular vector loadings. Significance testing for clinical variables relies on confidence intervals (significant if 0 ∉ [confidence interval]). Significance of imaging variables is based on calculating bootstrap ratios. Bootstrap ratios are computed by dividing the empirical singular vector loading by its bootstrapped standard error. A variable with a great contribution to the clinical-imaging relationship is characterized by a high singular vector loading which remains stable across bootstraps reflected by a small standard error. A bootstrap ratio was considered significant if it exceeded 1.96 or was lower than -1.96.

### Results

#### Text S8: Sample selection procedure

The UKB is an ongoing, multicenter, prospective population- based cohort study that enrolled 500.000 citizens, aged 40-69 years, from across the United Kingdom. The study collects in-depth genetic, physiological, lifestyle, environmental and imaging data. The multimodal imaging project aims to collect brain, heart, and abdomen scans from a subcohort of 100,000 participants. At the time of study initiation, 43098 individuals with brain MRIs were available. In addition to the internal quality assessment of the UKB further subject exclusion in the UKB cohort was performed based on the non-cancer illness codes (https://biobank.ndph.ox.ac.uk/showcase/coding.cgi?id=6). Excluded diseases included Alzheimer's disease, alcohol, opioid, and other addictions, amyotrophic lateral sclerosis, cerebral infarction, brain abscess, chronic neurological problem, encephalitis, epilepsy, haemorrhage, head injury, meningitis, multiple sclerosis, Parkinson's disease and skull fractures. Furthermore, individuals with PSMD values exceeding three standard deviations from the mean were excluded from the analysis. Subsequently, 38082 subjects were included into the following statistical analysis.

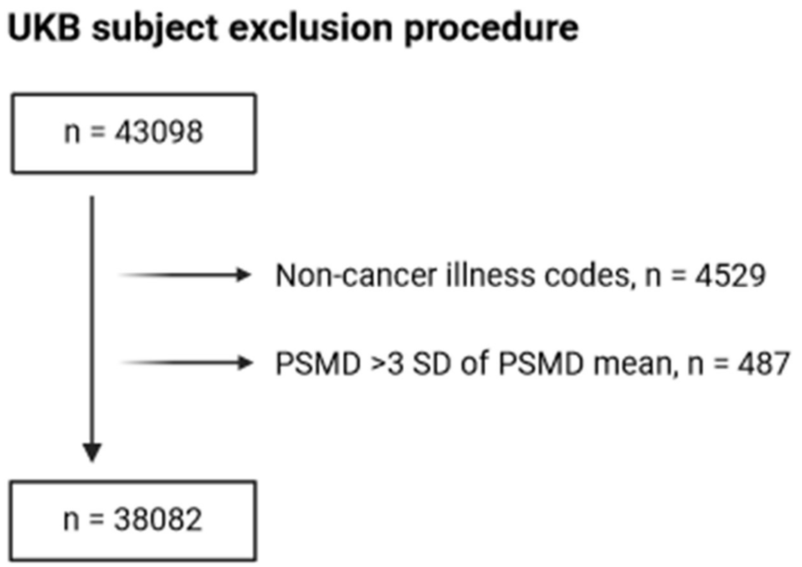

Subject exclusion chart UKB: Abbreviations: SD = Standard Deviation

The Hamburg City Health Study is an ongoing, monocentric, prospective, population-based cohort study that aims to explore prevalence, risk and prognostic factors of major chronic diseases. A random sample of 45000 persons aged 45-74 years from the general population of Hamburg is in the process of recruitment. Study participants undergo an extensive baseline examination, imaging and genomic, proteomic characterization [31]. At the stage of the present study, 2652 MRIs were available from the first 10000 study participants. Matching criteria to those used in the UKB were applied to HCHS subjects based on the neuroradiological evaluation and self-reported diagnoses. Additionally, subject exclusion in the HCHS was determined by issues encountered during MRI acquisition and preprocessing. In the UKB, this aspect had already been addressed through internal quality assessment procedures. In the process n = 155 subjects were excluded. Consequently, 2497 subjects served as study cohort for the statistical analysis.

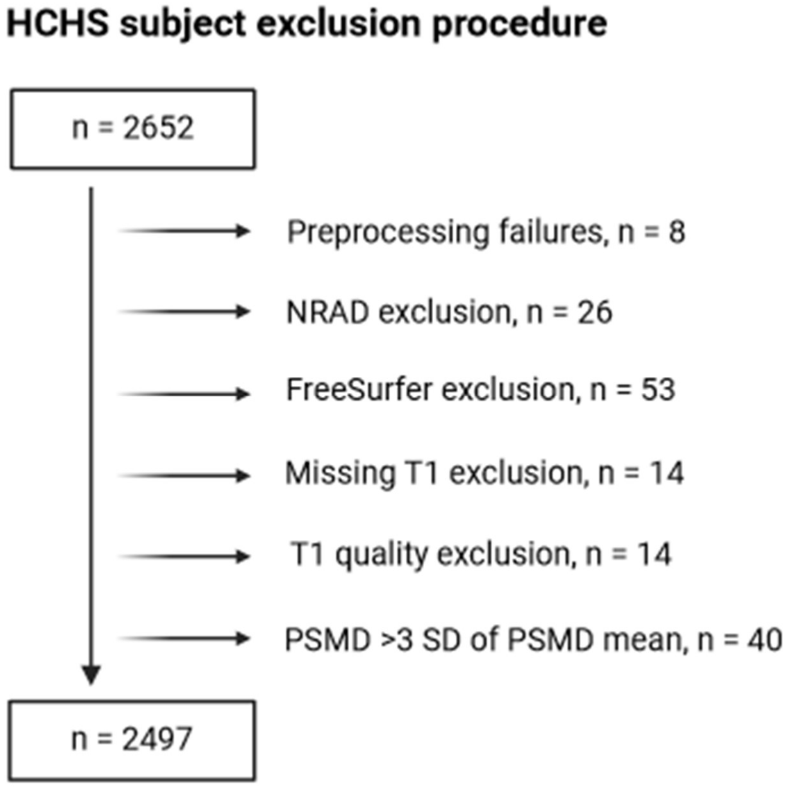

Subject exclusion chart HCHS: Abbreviations: SD = Standard Deviation

### Table S9: Partial least squares correlation results – UKB

| Clinical variables | Loading | Confidence interval |
|---|---|---|
| Executive function | $-8.0*10^{-2}$ | -0.07, -0.09 |
| Processing speed | $-9.1*10^{-2}$ | -0.08, -0.1 |
| Memory | $-5.4*10^{-2}$ | -0.04, -0.06 |
| Reasoning | $-7.1*10^{-2}$ | -0.06, -0.08 |
| Hand grip strength | $-2.8*10^{-2}$ | -0.02, -0.04 |
| Average acceleration | $-1.8*10^{-2}$ | -0.008, -0.03 |
| **Covariates** | **Loading** | **Confidence interval** |
| Age | $-1.3*10^{-17}$ | 0.01, -0.01 |
| Sex | $-1.0*10^{-17}$ | 0.01, -0.01 |
| Education | $1.4*10^{-18}$ | 0.01, -0.01 |

| Imaging variables | Bootstrap ratio |
|---|---|
| WMH | 12.4 |
| PSMD | 12.8 |
| Relative brain age | 19.8 |

### Table S10: Partial least squares correlation cross-validation – UKB

| **Cross-validation fold** | **Spearman correlation coefficient ($r_{sp}$)** | **$P_{FDR}$** |
|---|---|---|
| 1 | 0.084 | <0.001 |
| 2 | 0.074 | <0.001 |
| 3 | 0.100 | <0.001 |
| 4 | 0.093 | <0.001 |
| 5 | 0.103 | <0.001 |
| 6 | 0.090 | <0.001 |
| 7 | 0.114 | <0.001 |
| 8 | 0.096 | <0.001 |
| 9 | 0.081 | <0.001 |
| 10 | 0.127 | <0.001 |

PLS results of 10-fold cross-validation. Mean $r_{sp}$ = 0.096

### Figure S11: Partial least squares correlation including WMH load and PSMD brain age gap measures – UKB

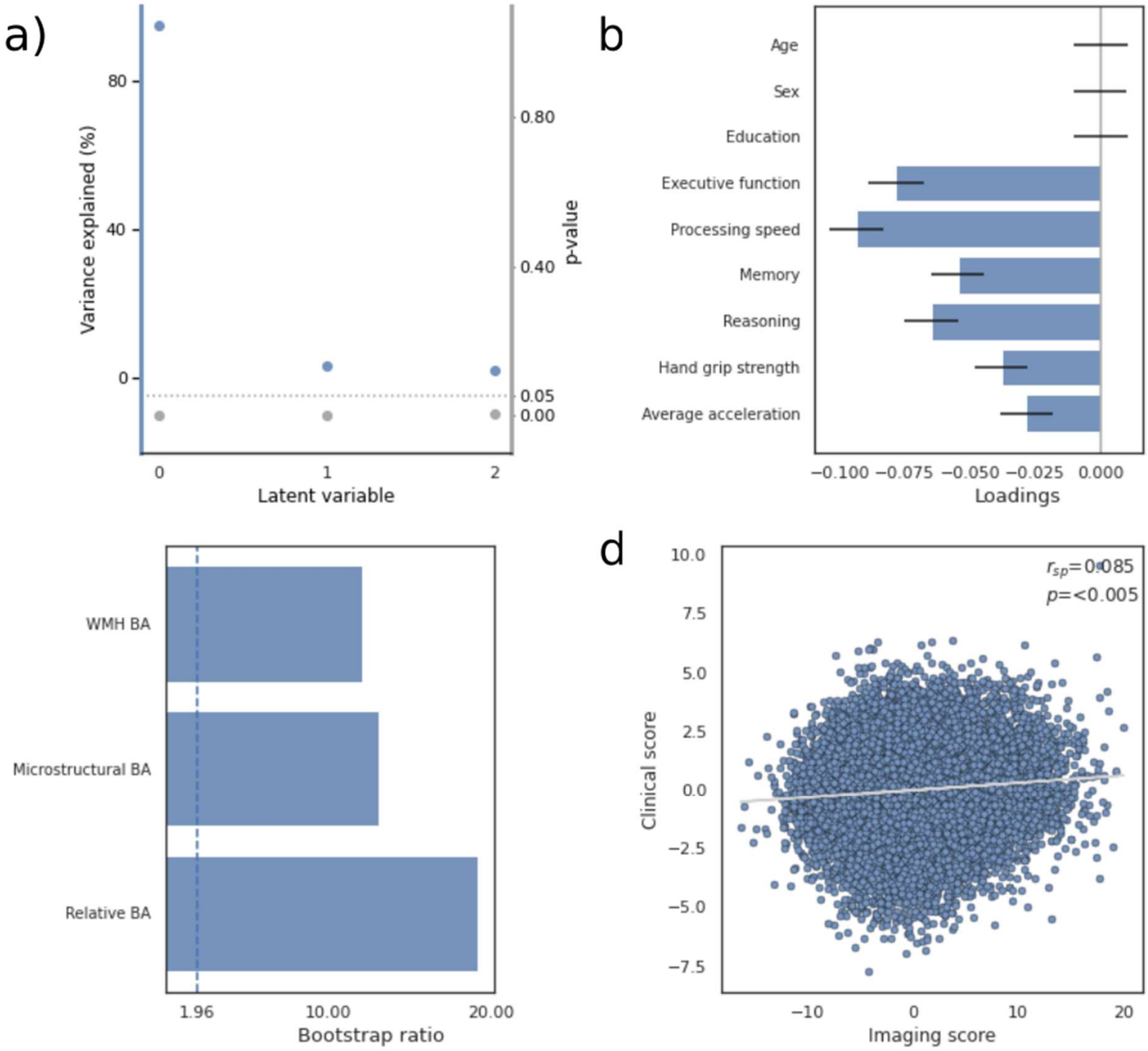

a) Overview of detected latent variables with the first latent variables explaining 94.6% of observed shared variance. b) Loadings of the clinical variable set consisting of cognitive and motor test results. Error bars indicate the 95% confidence interval obtained by bootstrap resampling. c) Bootstrap ratios of the imaging variable set. The vertical dashed line represents the significance threshold (bootstrap ratio > 1.96). d) Relationship of subject-level imaging and clinical scores. Abbreviations: $r_{sp}$ – Spearman correlation, WMH BA – WMH brain age; brain age gap measure based on logarithmic white matter hyperintensity load, microstructural BA – brain age gap measure based on peak width of skeletonized mean diffusivity, relative BA – relative brain age gap measure based on regional brain morphology.

#### Figure S12: Partial least squares correlation based on original cognitive and motor tests – UKB

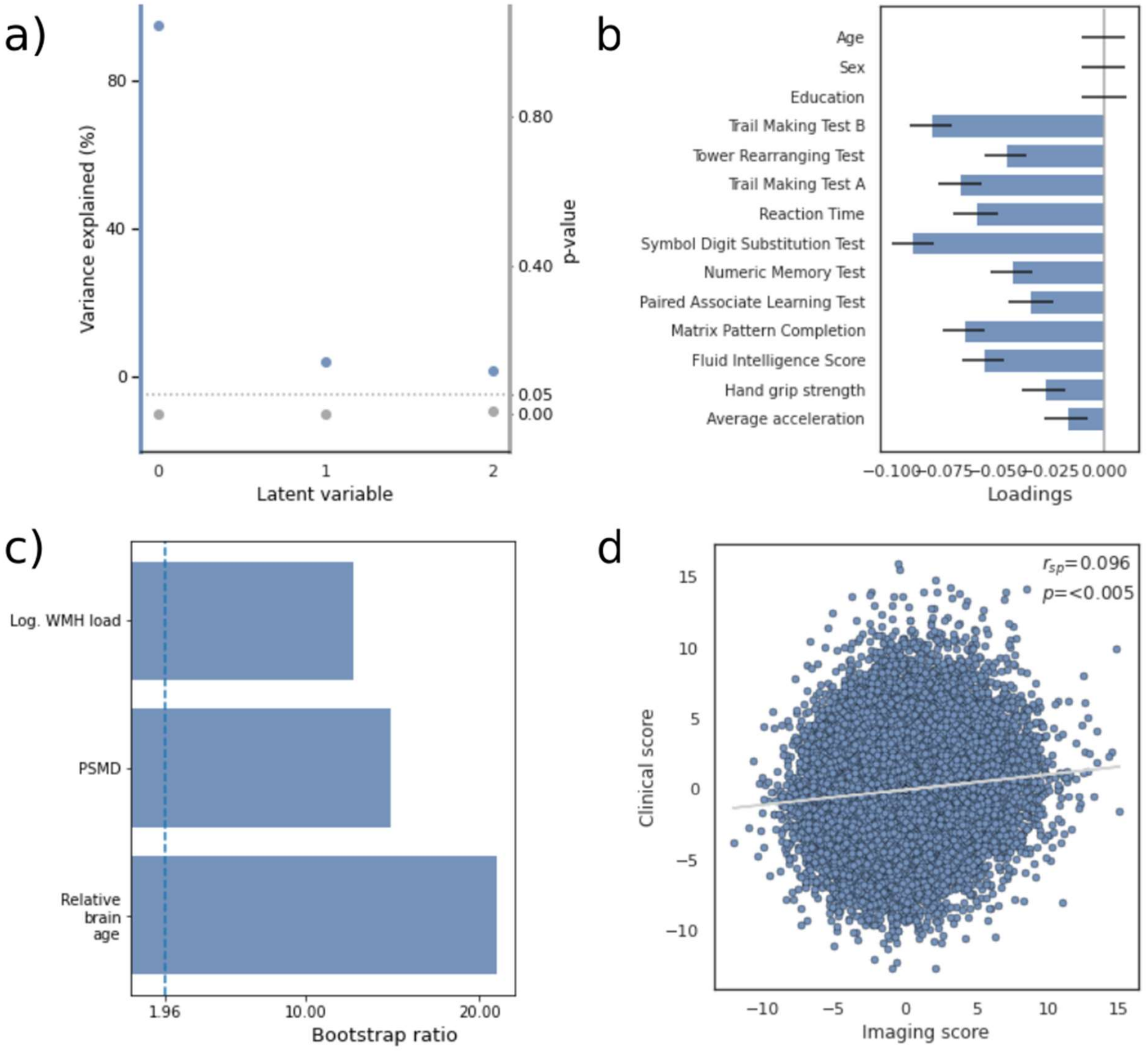

a) Overview of detected latent variables with the first latent variables explaining 94.6% of observed shared variance. b) Loadings of the clinical variable set consisting of cognitive and motor test results. Error bars indicate the 95% confidence interval obtained by bootstrap resampling. c) Bootstrap ratios of the imaging variable set. The vertical dashed line represents the significance threshold (bootstrap ratio > 1.96). d) Relationship of subject-level imaging and clinical scores. Please note that the Trail Making Test and reaction time results were reversed to align higher values with improved performance, consistent with the interpretation for the other cognitive test scores. Abbreviations: $r_{sp}$ – Spearman correlation, Log. WMH load – logarithmized white matter hyperintensity load, PSMD – Peak Width of Skeletonized Mean Diffusivity.

#### Table S13: Linear regression of imaging markers and cognitive/motor functions - UKB

| **Imaging - marker** / **Clinical marker** | WMH | | PSMD | | Relative brain age | |
|---|---|---|---|---|---|---|
| | $\beta_{std}$ | $P_{FDR}$ | $\beta_{std}$ | $P_{FDR}$ | $\beta_{std}$ | $P_{FDR}$ |
| Executive function | -0.05 | <0.001 | -0.05 | <0.001 | -0.07 | <0.001 |
| Processing speed | -0.05 | <0.001 | -0.08 | <0.001 | -0.08 | <0.001 |
| Memory | -0.06 | <0.001 | -0.03 | <0.001 | -0.05 | <0.001 |
| Reasoning | -0.06 | <0.001 | -0.01 | >0.05 | -0.07 | <0.001 |
| Hand grip strength | -0.03 | <0.001 | -0.03 | <0.001 | -0.02 | <0.001 |
| Average acceleration | -0.06 | <0.001 | -0.05 | <0.001 | -0.02 | 0.008 |

**Abbreviations:** $\beta_{std}$ = standardized β-coefficient of the respective regression model. $P_{FDR}$ = false discovery rate-corrected p-value.

#### Figure S14: Linear regression of imaging markers of biological brain age and cognitive/motor functions – UKB

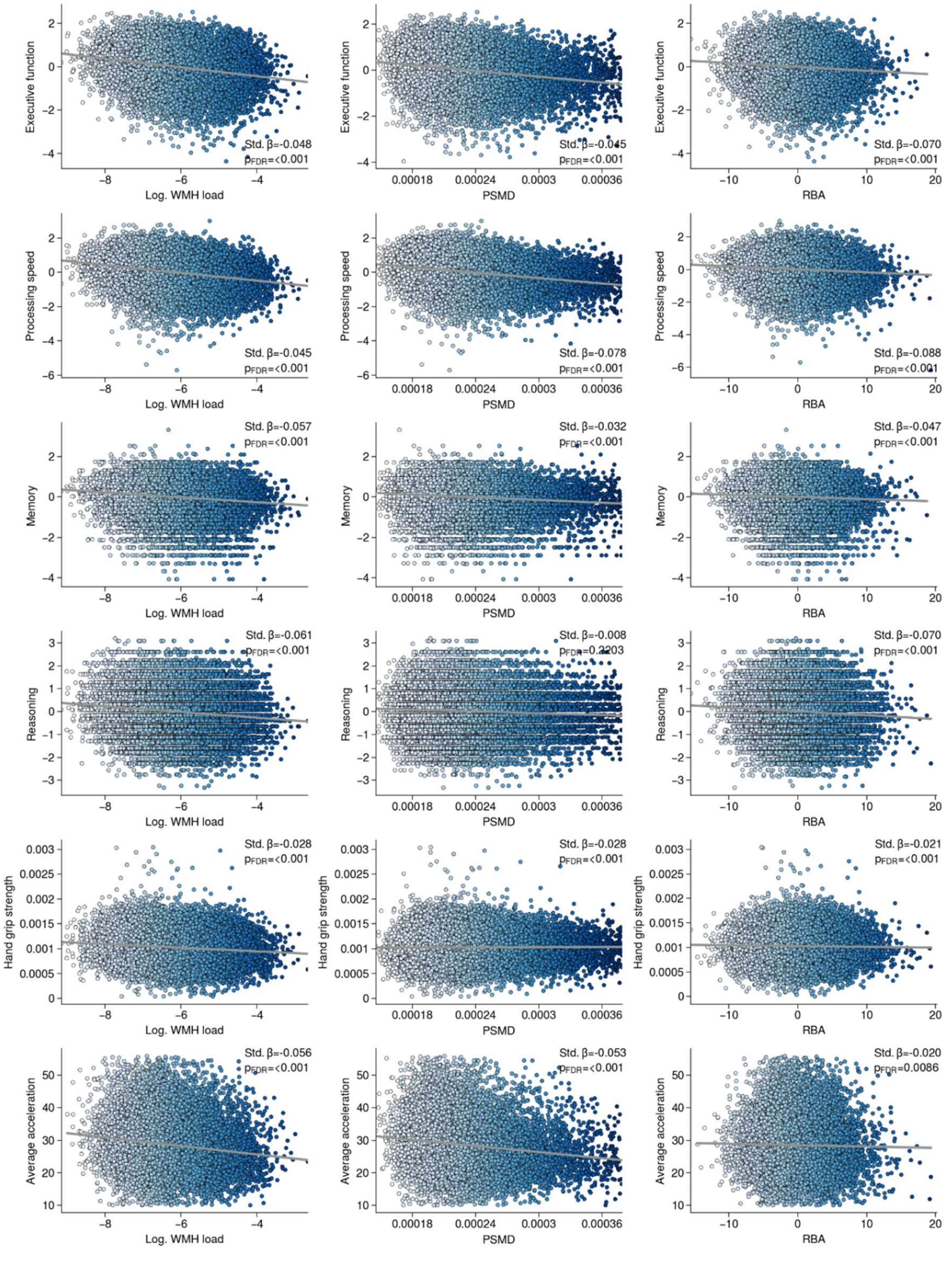

Regression plots display the association between imaging markers and cognitive and motor functions. All models were adjusted for age, sex and education. Effect sizes are presented as standardized β-coefficient (Std. β). p-values were false discovery rate-corrected ($P_{FDR}$).

### Figure S15: Cross correlation matrix of imaging markers - UKB

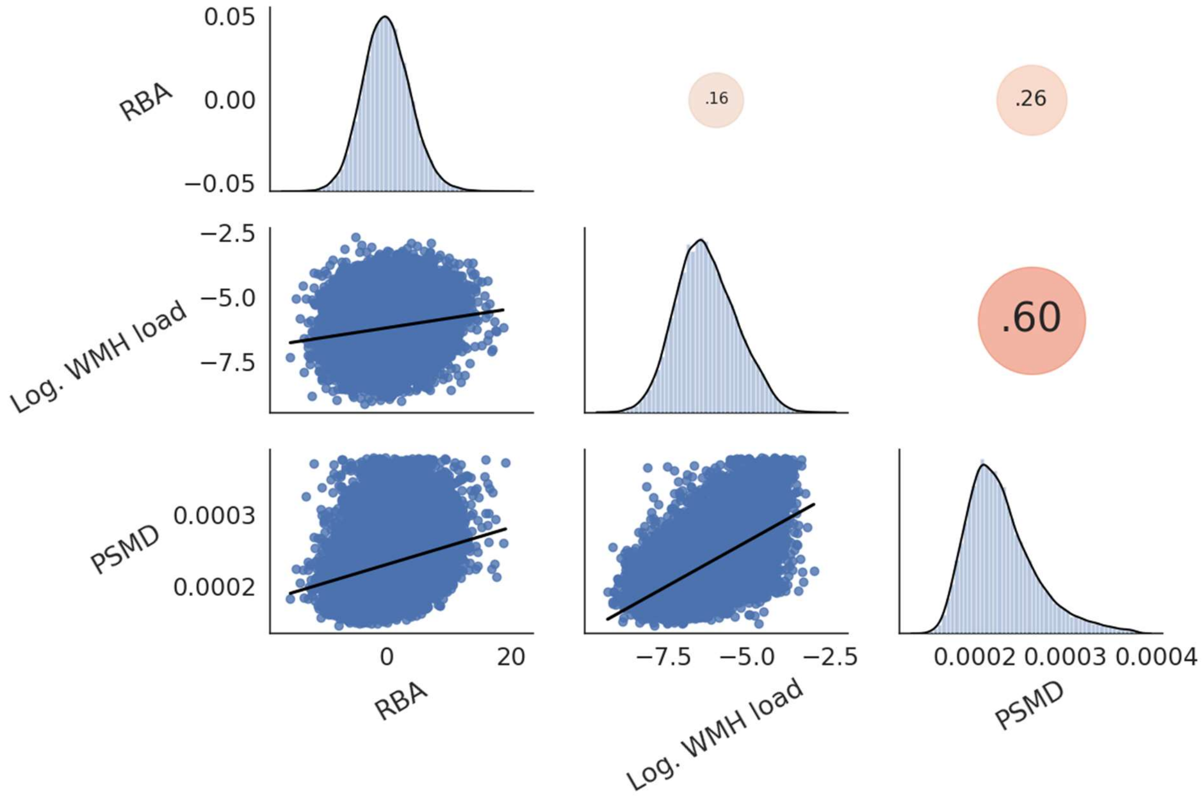

Regression plots in the lower triangle depict the linear association between imaging markers. In the upper triangle, dots represent the Pearson coefficient, with degree and direction illustrated by dot size and color (red indicating positive correlation, blue indicating negative correlation). Diagonal histograms illustrate the distribution of the respective imaging marker. Abbreviations: RBA – relative brain age, Log. WMH load – logarithmic white matter hyperintensity load, PSMD – peak width of skeletonized mean diffusivity.

#### Figure S16: Cross correlation matrix of clinical markers - UKB

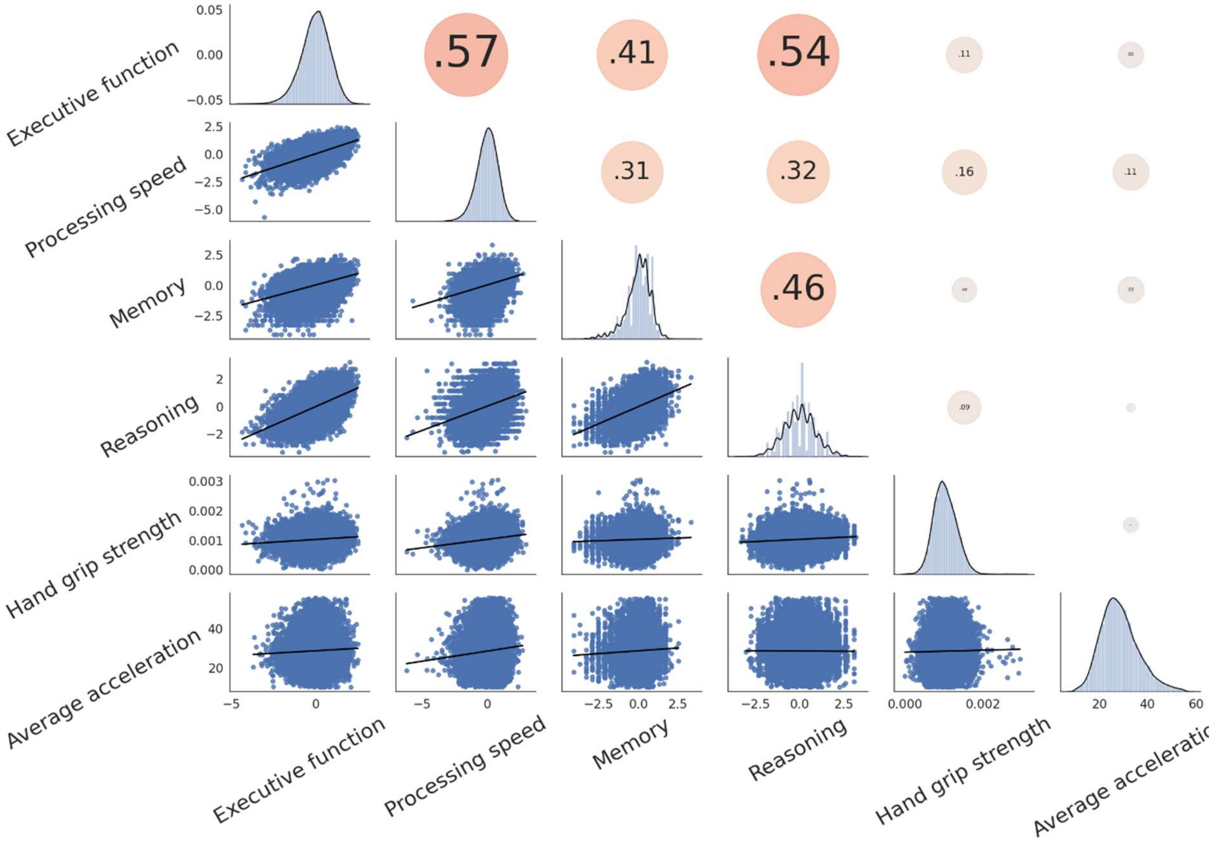

Regression plots in the lower triangle depict the linear association between cognitive and motor scores. In the upper triangle, dots represent the Pearson correlation coefficient, with degree and direction illustrated by dot size and color (red indicating positive correlation, blue indicating negative correlation). Diagonal histograms illustrate the distribution of the respective clinical marker.

#### Figure S17: Partial least squares correlation - HCHS

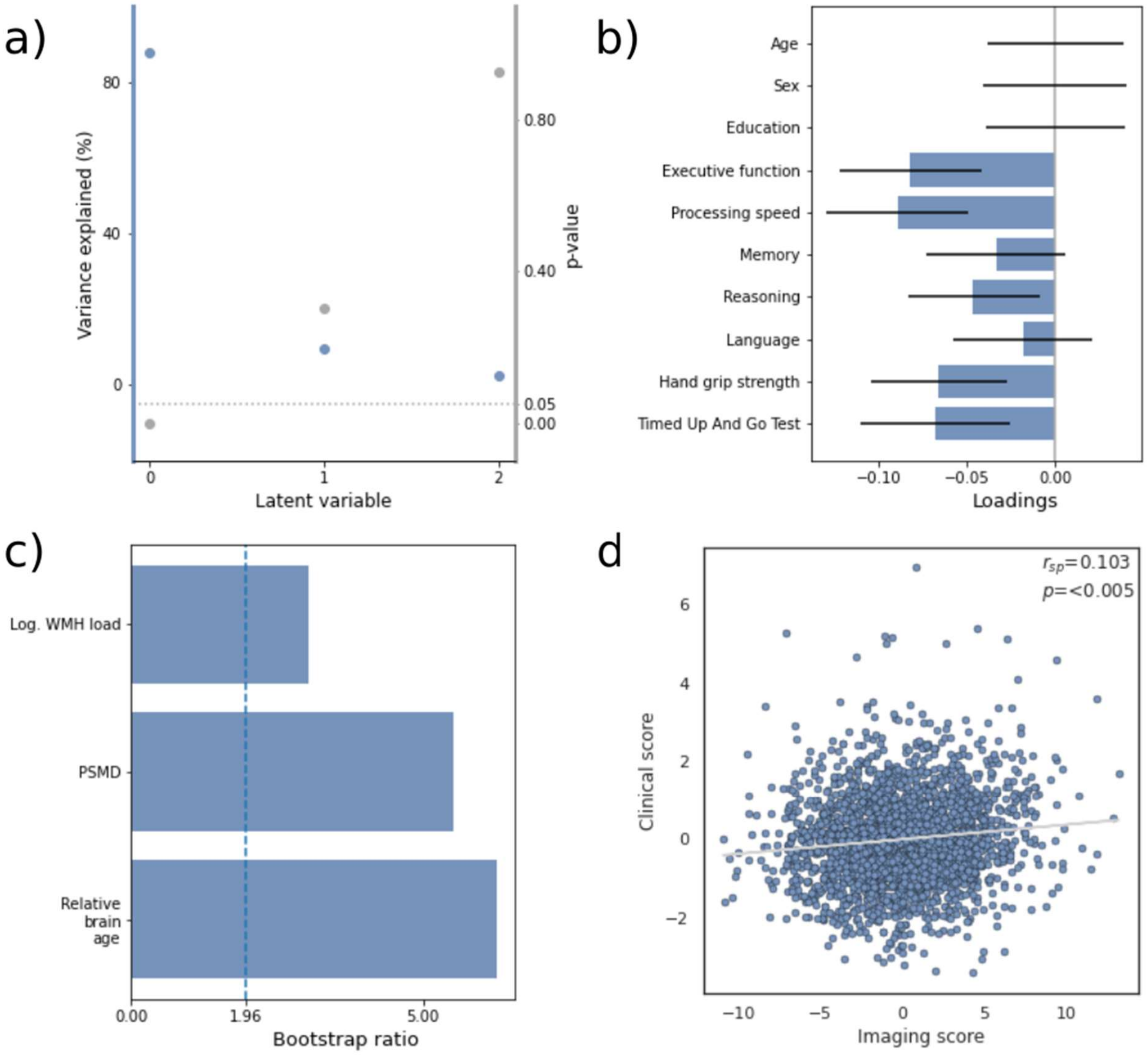

a) Overview of detected latent variables with the first latent variables explaining 87.7% of observed shared variance. b) Loadings of the clinical variable set consisting of cognitive and motor test results. Error bars indicate the 95% confidence interval obtained by bootstrap resampling. c) Bootstrap ratios of the imaging variable set. The vertical dashed line represents the significance threshold (bootstrap ratio > 1.96). d) Relationship of subject-level imaging and clinical scores. Please note that the Timed Up And Go Test results were reversed before PLS to align higher values with improved performance, consistent with the interpretation for the other test scores. Abbreviations: $r_{sp}$ – Spearman correlation, Log. WMH load – logarithmized white matter hyperintensity load, PSMD – Peak Width of Skeletonized Mean Diffusivity.

### Table S18: Partial least squares correlation - HCHS

| Clinical variables | Loading | Confidence interval |
|---|---|---|
| Executive function | $-8.2*10^{-2}$ | -0.04, -0.12 |
| Processing speed | $-8.9*10^{-02}$ | -0.05, -0.13 |
| Memory | $-3.3*10^{-02}$ | 0.01, -0.07 |
| Reasoning | $-4.7*10^{-02}$ | -0.01, -0.08 |
| Language | $-1.8*10^{-02}$ | 0.02, -0.06 |
| Hand grip strength | $-6.6*10^{-02}$ | -0.03, -0.10 |
| Timed Up And Go Test | $-6.7*10^{-02}$ | -0.02, -0.11 |
| **Covariates** | **Loading** | **Confidence interval** |
| Age | $1.1*10^{-17}$ | 0.04, -0.04 |
| Sex | $-1.3*10^{-17}$ | 0.04, -0.04 |
| Education | $-1.5*10^{-17}$ | 0.04, -0.04 |

| Imaging variables | Bootstrap ratio |
|---|---|
| WMH | 3.0 |
| PSMD | 5.5 |
| Relative brain age | 6.3 |

### Table S19: Partial least squares correlation cross-validation – HCHS

| **Cross-validation fold** | **Spearman correlation coefficient ($r_{sp}$)** | **$P_{FDR}$** |
|---|---|---|
| 1 | 0.115 | >0.05 |
| 2 | 0.079 | >0.05 |
| 3 | 0.126 | >0.05 |
| 4 | 0.121 | >0.05 |
| 5 | -0.018 | >0.05 |
| 6 | 0.125 | >0.05 |
| 7 | 0.219 | 0.005 |
| 8 | 0.137 | >0.05 |
| 9 | 0.027 | >0.05 |
| 10 | 0.144 | >0.05 |

PLS results validated via 10-fold cross-validation. Mean $r_{sp}$ = 0.108

### Figure S20: Partial least squares correlation including WMH load and PSMD brain age gap measures – HCHS

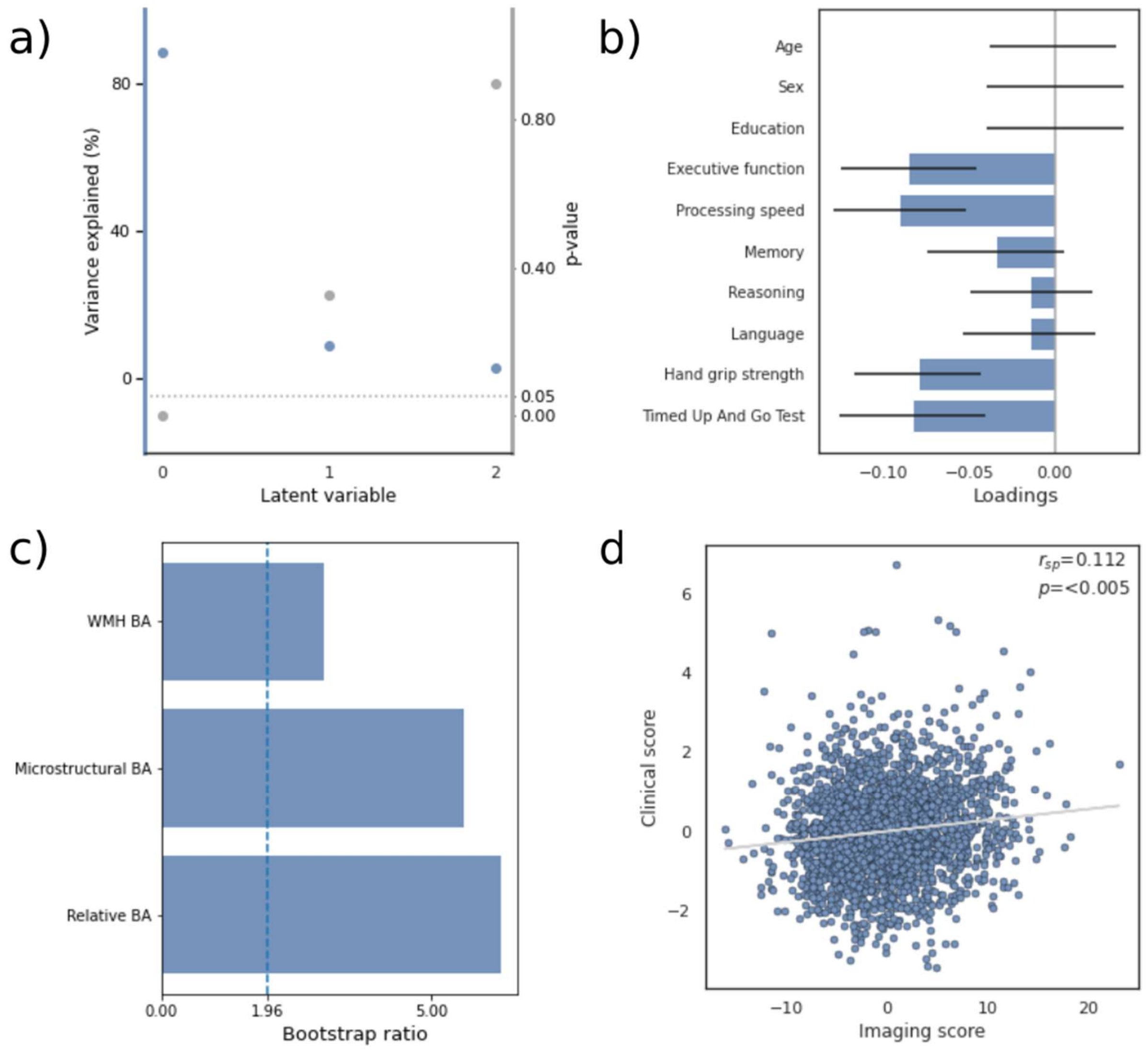

a) Overview of detected latent variables with the first latent variables explaining 88.2% of observed shared variance. b) Loadings of the clinical variable set consisting of cognitive and motor test results. Error bars indicate the 95% confidence interval obtained by bootstrap resampling. c) Bootstrap ratios of the imaging variable set. The vertical dashed line represents the significance threshold (bootstrap ratio > 1.96). d) Relationship of subject-level imaging and clinical scores. Please note that the Timed Up And Go Test results were reversed before PLS to align higher values with improved performance, consistent with the interpretation for the other test scores. Abbreviations: $r_{sp}$ – Spearman correlation, WMH BA – WMH brain age; brain age gap measure based on logarithmic white matter hyperintensity load, microstructural BA – brain age gap measure based on peak width of skeletonized mean diffusivity, Relative BA – relative brain age gap measure based on regional brain morphology.

### Figure S21: Partial least squares correlation based on original cognitive and motor tests – HCHS

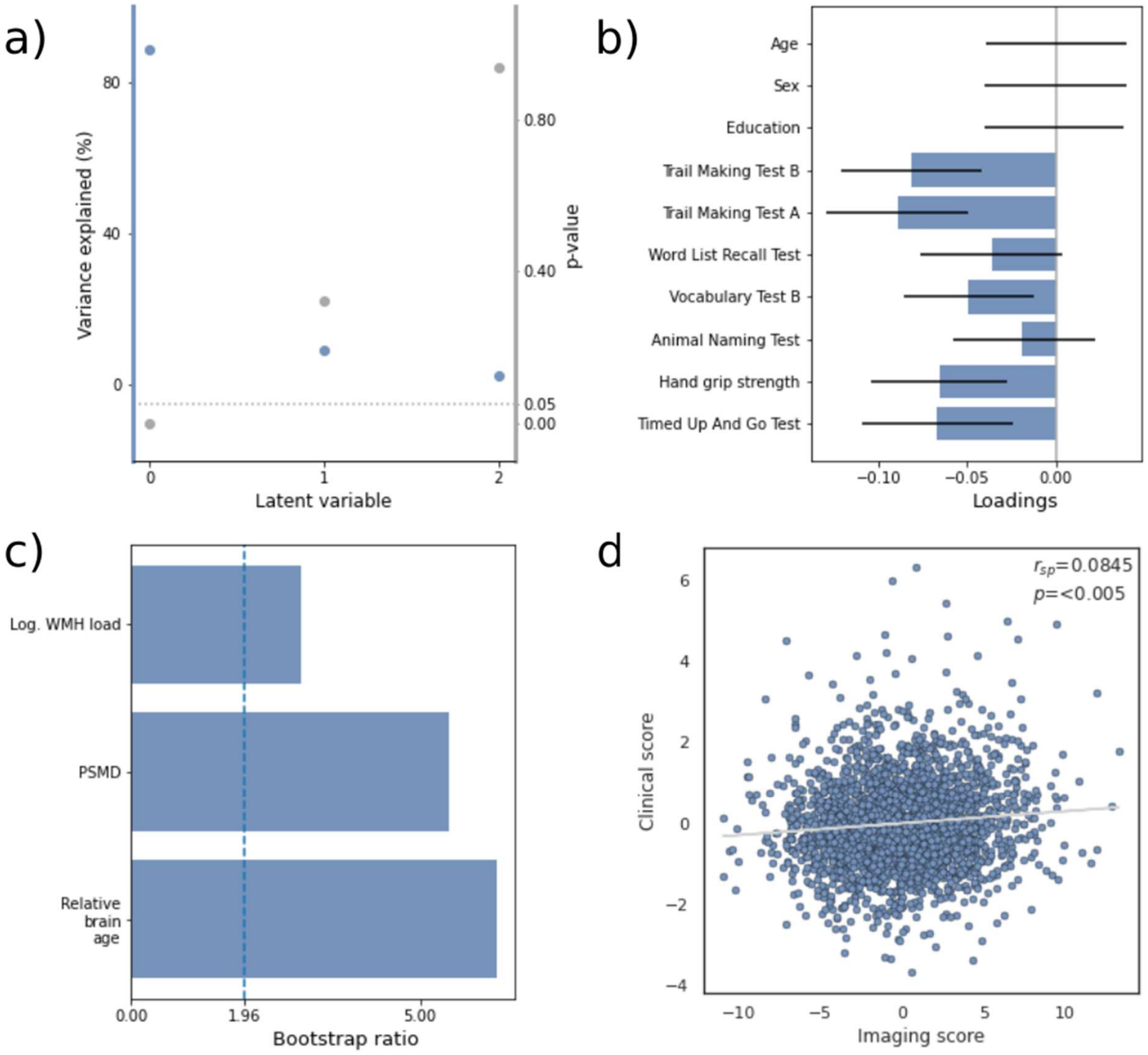

a) Overview of detected latent variables with the first latent variables explaining 88.3% of observed shared variance. b) Loadings of the clinical variable set consisting of cognitive and motor test results. Error bars indicate the 95% confidence interval obtained by bootstrap resampling. c) Bootstrap ratios of the imaging variable set. The vertical dashed line represents the significance threshold (bootstrap ratio > 1.96). d) Relationship of subject-level imaging and clinical scores. Please note that the Trail Making Test and Timed Up And Go Test results were reversed to align higher values with improved performance, consistent with the interpretation for the other test scores. Abbreviations: $r_{sp}$ – Spearman correlation, Log. WMH load – logarithmized white matter hyperintensity load, PSMD – Peak Width of Skeletonized Mean Diffusivity.

#### Table S22: Linear regression of imaging markers and cognitive/motor functions - HCHS

| Imaging marker / Clinical marker | WMH | | PSMD | | Relative brain age | |
|---|---|---|---|---|---|---|
| | $\beta_{std}$ | $P_{FDR}$ | $\beta_{std}$ | $P_{FDR}$ | $\beta_{std}$ | $P_{FDR}$ |
| Executive function | -0.057 | >0.05 | -0.052 | >0.05 | -0.071 | <0.001 |
| Processing speed | -0.043 | >0.05 | -0.070 | 0.013 | -0.079 | <0.001 |
| Memory | -0.001 | >0.05 | 0.008 | >0.05 | -0.034 | >0.05 |
| Reasoning | -0.006 | >0.05 | 0.027 | >0.05 | -0.053 | 0.016 |
| Language | 0.019 | >0.05 | -0.018 | >0.05 | -0.018 | >0.05 |
| Hand grip strength | -0.010 | >0.05 | -0.070 | 0.002 | -0.054 | 0.002 |
| Timed Up And Go Test | -0.027 | >0.05 | -0.107 | 0.002 | -0.079 | 0.002 |

Abbreviations: $\beta_{std}$ = standardized β-coefficient of the respective regression model. $P_{FDR}$ = false discovery rate-corrected p-value.

### Figure S23: Linear regression of imaging markers and cognitive/motor function

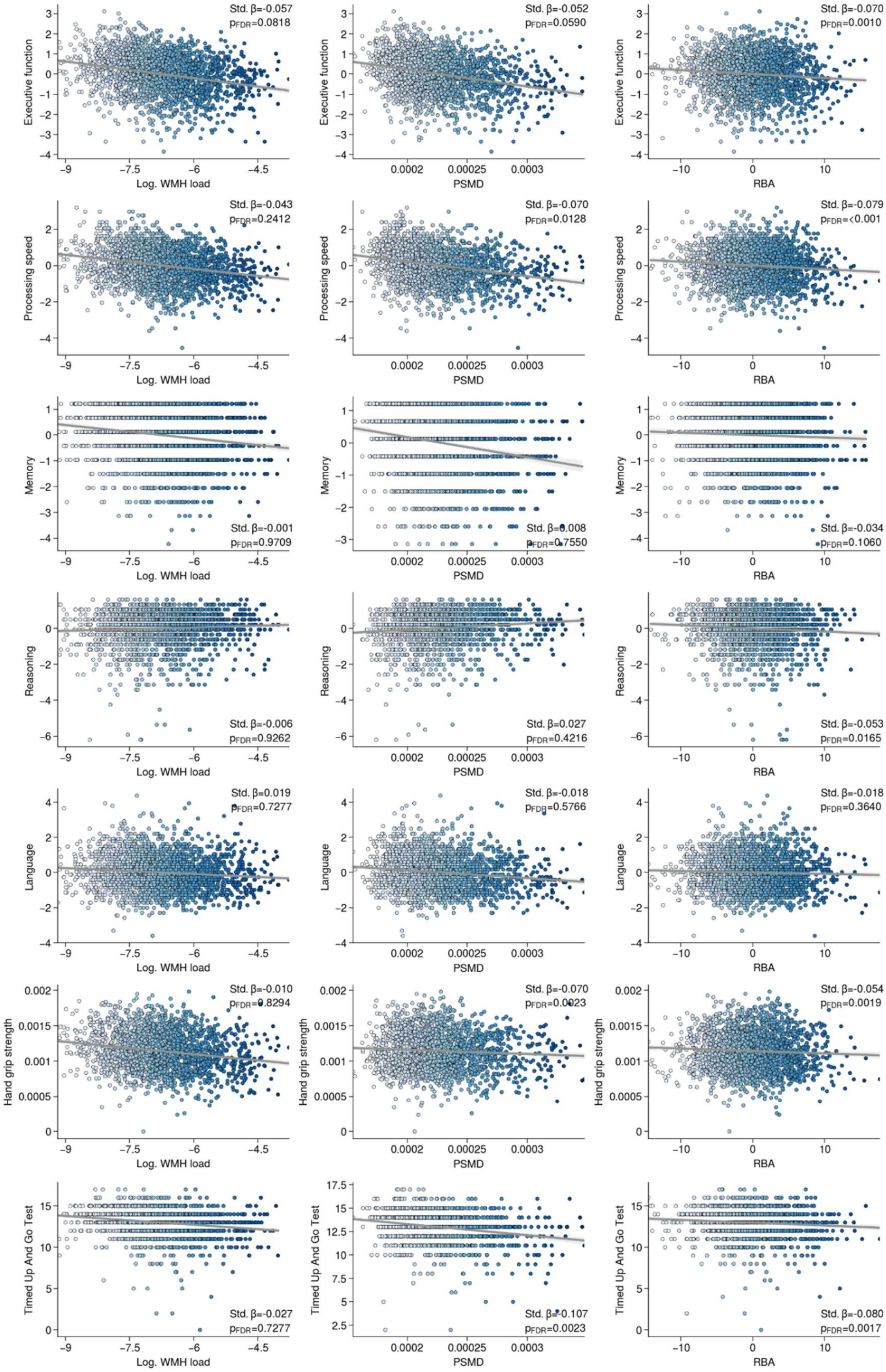

Regression plots illustrate the association between imaging markers and cognitive and motor functions. All models were adjusted for age, sex and education. Effect sizes are presented as standardized β-coefficient (Std. β). p-values were false discovery rate-corrected ($P_{FDR}$).

#### Figure S24: Cross correlation matrix of imaging markers - HCHS

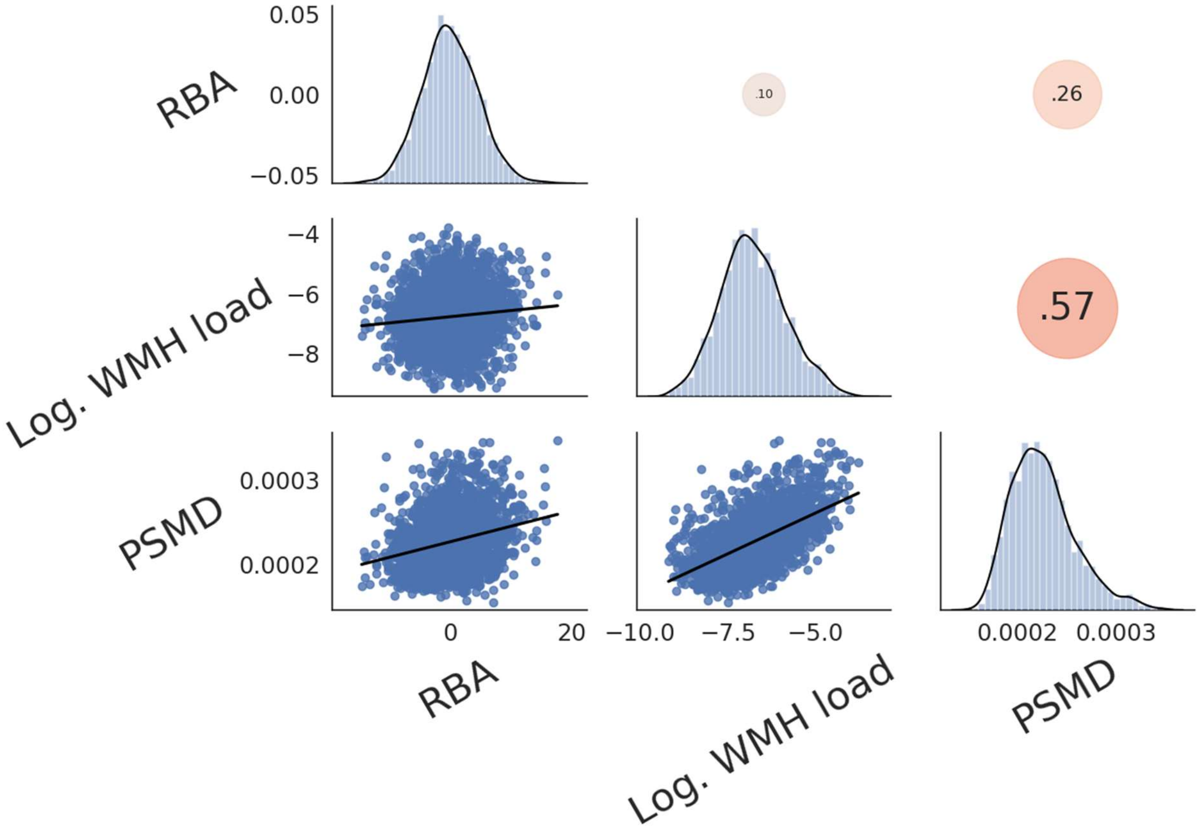

Regression plots in the lower triangle depict the linear association between imaging markers. In the upper triangle, dots represent the Pearson correlation coefficient, with degree and direction depicted by dot size and color (red indicating positive correlation, blue indicating negative correlation). Diagonal histograms illustrate the distribution of the respective imaging markers. Abbreviations: RBA – relative brain age, Log. WMH load - logarithmic white matter hyperintensity load, PSMD - peak width of skeletonized mean diffusivity.

### Figure S25: Cross correlation matrix of clinical markers - HCHS

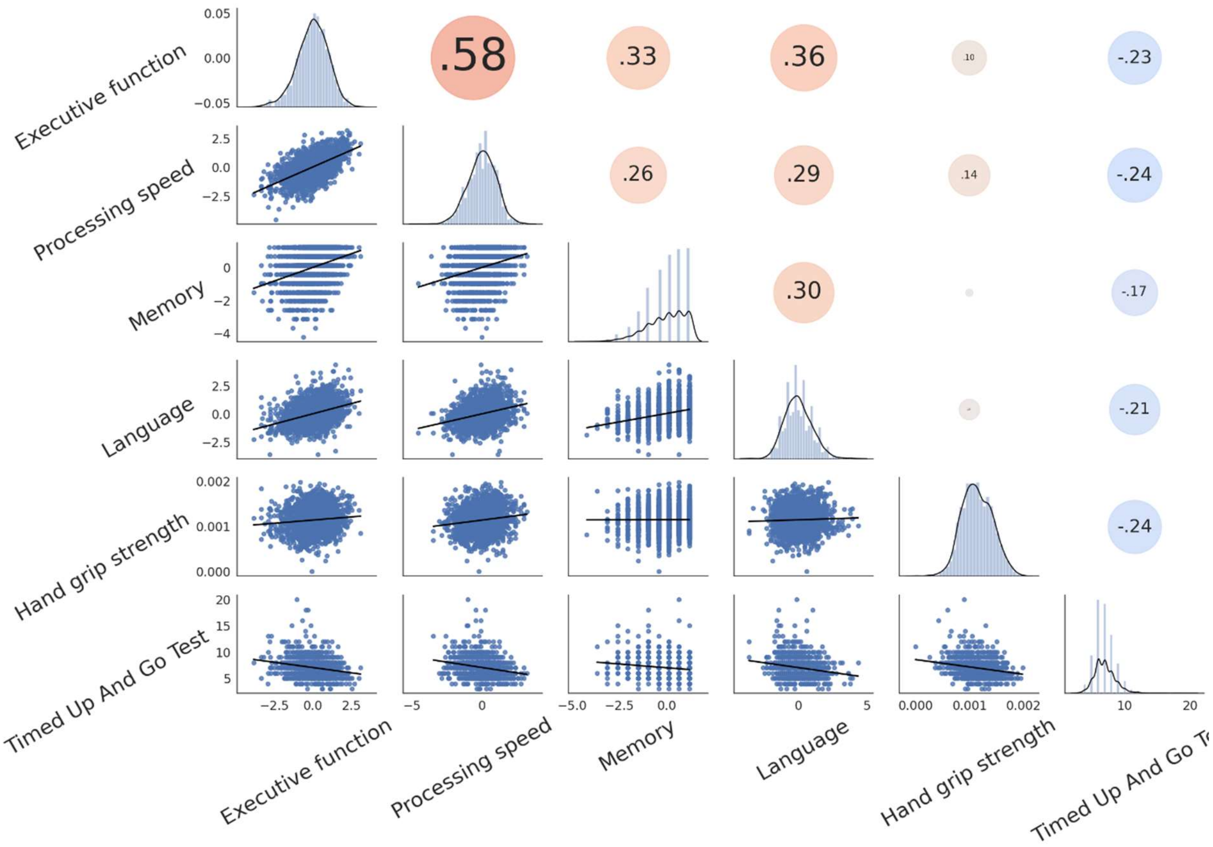

Regression plots in the lower triangle depict the linear association between cognitive and motor scores. In the upper triangle, dots represent the Pearson coefficient, with degree and direction depicted by dot size and color (red indicating positive correlation, blue indicating negative correlation). Diagonal histograms illustrate the distribution of the respective clinical marker.

### Figure S26: Mediation analysis results - HCHS

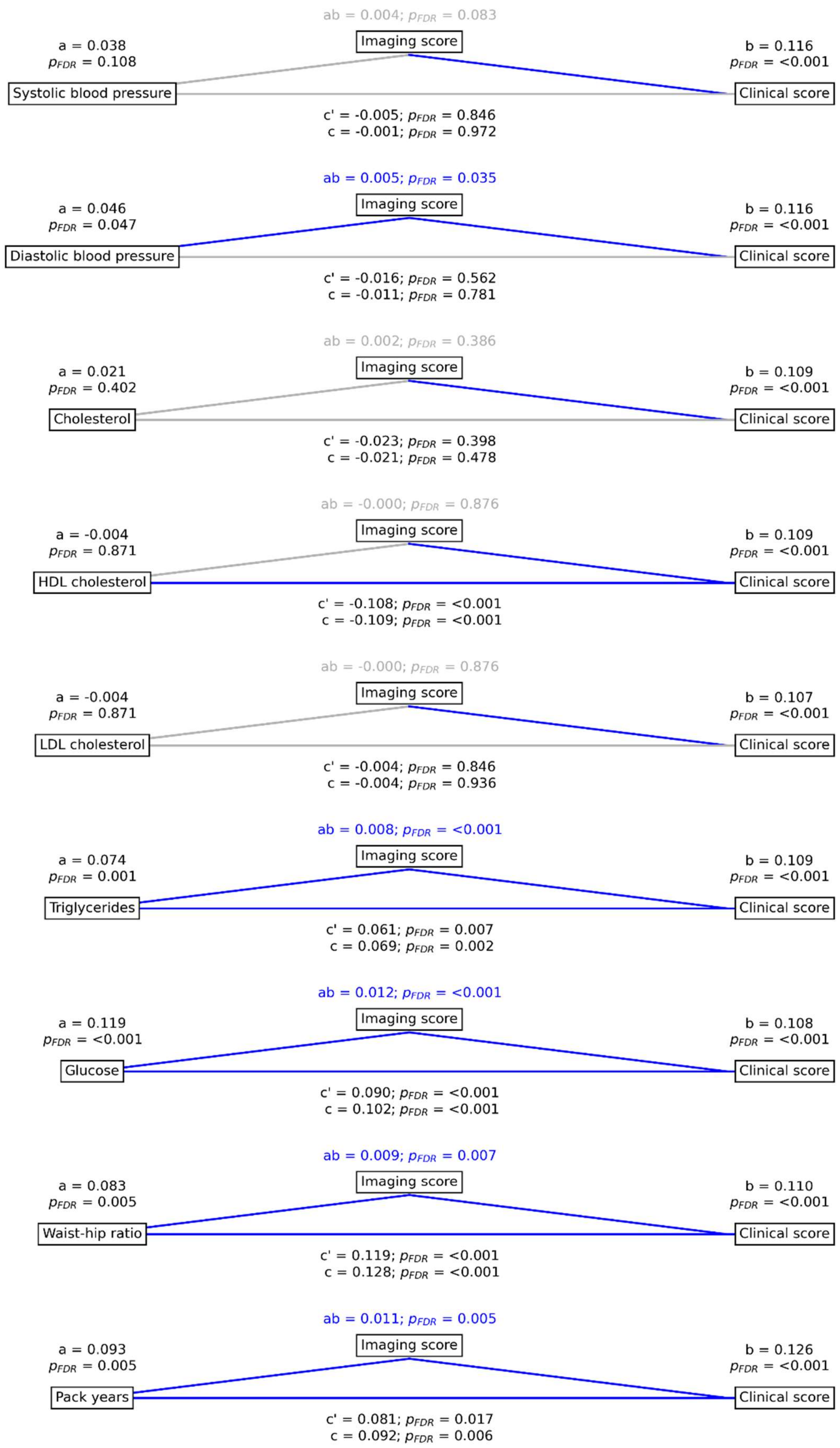

Mediation analysis results of the HCHS. Mediation effects of the subject-level imaging score on the relationship between vascular risk factors and the subject-level clinical score summarizing cognitive and motor performance. Path plots display standardized effects and p-values: (a) vascular risk factor to imaging score, (b) subject-level imaging score to clinical score, (ab) indirect effect (c') direct effect and (c) total effect. Significant paths are highlighted in blue; non-significant in light gray. If the indirect path was significant the text for ab is highlighted in blue. Abbreviations: $P_{FDR}$ – false discovery rate-corrected p-value.

### References

1. Doherty A, Jackson D, Hammerla N, et al. Large Scale Population Assessment of Physical Activity Using Wrist Worn Accelerometers: The UK Biobank Study. PloS One. 2017;12(2):e0169649. doi:10.1371/journal.pone.0169649

2. Fawns-Ritchie C, Deary IJ. Reliability and validity of the UK Biobank cognitive tests. PloS One. 2020;15(4):e0231627. doi:10.1371/journal.pone.0231627

3. Podsiadlo D, Richardson S. The timed “Up & Go”: a test of basic functional mobility for frail elderly persons. J Am Geriatr Soc. 1991;39(2):142-148. doi:10.1111/j.1532-5415.1991.tb01616.x

4. Campagna F, Montagnese S, Ridola L, et al. The animal naming test: An easy tool for the assessment of hepatic encephalopathy. Hepatol Baltim Md. 2017;66(1):198-208. doi:10.1002/hep.29146

5. Tombaugh TN. Trail Making Test A and B: normative data stratified by age and education. Arch Clin Neuropsychol Off J Natl Acad Neuropsychol. 2004;19(2):203-214. doi:10.1016/S0887-6177(03)00039-8

6. Morris JC, Heyman A, Mohs RC, et al. The Consortium to Establish a Registry for Alzheimer’s Disease (CERAD). Part I. Clinical and neuropsychological assessment of Alzheimer’s disease. Neurology. 1989;39(9):1159-1165. doi:10.1212/wnl.39.9.1159

7. Miller KL, Alfaro-Almagro F, Bangerter NK, et al. Multimodal population brain imaging in the UK Biobank prospective epidemiological study. Nat Neurosci. 2016;19(11):1523-1536. doi:10.1038/nn.4393

8. Petersen M, Frey BM, Schlemm E, et al. Network Localisation of White Matter Damage in Cerebral Small Vessel Disease. Sci Rep. 2020;10(1):9210. doi:10.1038/s41598-020-66013-w

9. Petersen M, Frey BM, Schlemm E, et al. Network Localisation of White Matter Damage in Cerebral Small Vessel Disease. Sci Rep. 2020;10(1):9210. doi:10.1038/s41598-020-66013-w

10. Tustison NJ, Avants BB, Cook PA, et al. N4ITK: improved N3 bias correction. IEEE Trans Med Imaging. 2010;29(6):1310-1320. doi:10.1109/TMI.2010.2046908

11. Fischl B, Dale AM. Measuring the thickness of the human cerebral cortex from magnetic resonance images. Proc Natl Acad Sci. 2000;97(20):11050-11055. doi:10.1073/pnas.200033797

12. Fischl B, Salat DH, Busa E, et al. Whole brain segmentation: automated labeling of neuroanatomical structures in the human brain. Neuron. 2002;33(3):341-355. doi:10.1016/s0896-6273(02)00569-x

13. Dale AM, Fischl B, Sereno MI. Cortical surface-based analysis. I. Segmentation and surface reconstruction. NeuroImage. 1999;9(2):179-194. doi:10.1006/nimg.1998.0395

14. Cieslak M, Cook PA, He X, et al. QSIPrep: an integrative platform for preprocessing and reconstructing diffusion MRI data. Nat Methods. 2021;18(7):775-778. doi:10.1038/s41592-021-01185-5

15. Veraart J, Novikov DS, Christiaens D, Ades-Aron B, Sijbers J, Fieremans E. Denoising of diffusion MRI using random matrix theory. NeuroImage. 2016;142:394-406. doi:10.1016/j.neuroimage.2016.08.016

16. Kellner E, Dhital B, Kiselev VG, Reisert M. Gibbs-ringing artifact removal based on local subvoxel-shifts. Magn Reson Med. 2016;76(5):1574-1581. doi:10.1002/mrm.26054

17. Andersson JLR, Sotiropoulos SN. An integrated approach to correction for off-resonance effects and subject movement in diffusion MR imaging. NeuroImage. 2016;125:1063-1078. doi:10.1016/j.neuroimage.2015.10.019

18. Esteban O, Markiewicz CJ, Blair RW, et al. fMRIPrep: a robust preprocessing pipeline for functional MRI. Nat Methods. 2019;16(1):111-116. doi:10.1038/s41592-018-0235-4

19. Huntenburg J, Str L. Evaluating nonlinear coregistration of BOLD EPI and T1w images.

20. Power JD, Mitra A, Laumann TO, Snyder AZ, Schlaggar BL, Petersen SE. Methods to detect, characterize, and remove motion artifact in resting state fMRI. NeuroImage. 2014;84:320-341. doi:10.1016/j.neuroimage.2013.08.048

21. Abraham A, Pedregosa F, Eickenberg M, et al. Machine learning for neuroimaging with scikit-learn. Front Neuroinformatics. 2014;8:14. doi:10.3389/fninf.2014.00014

22. Garyfallidis E, Brett M, Amirbekian B, et al. Dipy, a library for the analysis of diffusion MRI data. Front Neuroinformatics. 2014;8:8. doi:10.3389/fninf.2014.00008

23. Petersen M, Nägele FL, Mayer C, et al. Brain imaging and neuropsychological assessment of individuals recovered from a mild to moderate SARS-CoV-2 infection. Proc Natl Acad Sci. 2023;120(22):e2217232120. doi:10.1073/pnas.2217232120

24. Petersen M, Nägele FL, Mayer C, et al. Brain network architecture constrains age-related cortical thinning. NeuroImage. 2022;264:119721. doi:10.1016/j.neuroimage.2022.119721

25. Klapwijk ET, van de Kamp F, van der Meulen M, Peters S, Wierenga LM. Qoala-T: A supervised-learning tool for quality control of FreeSurfer segmented MRI data. NeuroImage. 2019;189:116-129. doi:10.1016/j.neuroimage.2019.01.014

26. Petersen M, Nägele FL, Mayer C, et al. Brain imaging and neuropsychological assessment of individuals recovered from mild to moderate SARS-CoV-2 infection. Published online July 9, 2022:2022.07.08.22277420. doi:10.1101/2022.07.08.22277420

27. Fischl B, Salat DH, Busa E, et al. Whole brain segmentation: automated labeling of neuroanatomical structures in the human brain. Neuron. 2002;33(3):341-355. doi:10.1016/s0896-6273(02)00569-x

28. Fischl B, Dale AM. Measuring the thickness of the human cerebral cortex from magnetic resonance images. Proc Natl Acad Sci U S A. 2000;97(20):11050-11055. doi:10.1073/pnas.200033797

29. Griffanti L, Zamboni G, Khan A, et al. BIANCA (Brain Intensity AbNormality Classification Algorithm): A new tool for automated segmentation of white matter hyperintensities. NeuroImage. 2016;141:191-205. doi:10.1016/j.neuroimage.2016.07.018

30. Sundaresan V, Zamboni G, Le Heron C, et al. Automated lesion segmentation with BIANCA: Impact of population-level features, classification algorithm and locally adaptive thresholding. NeuroImage. 2019;202:116056. doi:10.1016/j.neuroimage.2019.116056

31. Jagodzinski A, Johansen C, Koch-Gromus U, et al. Rationale and Design of the Hamburg City Health Study. Eur J Epidemiol. 2020;35(2):169-181. doi:10.1007/s10654-019-00577-4
